# Allelic Complexity of *KMT2A* Partial Tandem Duplications in Acute Myeloid Leukemia and Myelodysplastic Syndromes

**DOI:** 10.1101/2021.11.01.21265781

**Authors:** Harrison K. Tsai, Christopher J. Gibson, H. Moses Murdock, Phani Davineni, Marian H. Harris, Eunice S. Wang, Lukasz P. Gondek, Annette S. Kim, Valentina Nardi, R. Coleman Lindsley

**Author notes:** **Corresponding Author:** R. Coleman Lindsley, MD, PhD, Dana-Farber Cancer Institute, 450 Brookline Avenue, Boston, MA 02215.

## Abstract

*KMT2A* partial tandem duplication (*KMT2A*-PTD) at 11q23.3 is associated with adverse risk in AML and MDS, is a potential therapeutic target, and is an attractive marker of measurable residual disease. High initial *KMT2A*-PTD RNA levels have been linked to poor prognosis, but mechanisms regulating *KMT2A*-PTD expression are not well understood. While it has been reported that *KMT2A*-PTD affects only a single allele, it has been theorized but not proven that duplications or genomic gains of a monoallelic *KMT2A*-PTD may occur, thereby potentially driving high expression and disease progression. Copy neutral loss of heterozygosity (CN-LOH) of 11q has also been described and is known to be associated with mutations in *CBL* but has not been reported to involve *KMT2A*-PTD. In this study, we identified 94 patients with *KMT2A*-PTDs using targeted DNA next-generation sequencing (NGS) and found that 16% (15/94) had complex secondary events, including CN-LOH and selective gain involving the *KMT2A*-PTD allele. High copy numbers indicating complexity were significantly enriched in AML versus MDS and correlated with higher RNA expression. Moreover, in serial samples, complexity was associated with relapse and secondary transformation. Taken together, we provide approaches to integrate quantitative and allelic assessment of *KMT2A*-PTDs into targeted DNA NGS and demonstrate that secondary genetic events occur in *KMT2A*-PTD by multiple mechanisms that may be linked to myeloid disease progression by driving increased expression from the affected allele.

## Introduction

Partial tandem duplication (PTD) of the lysine methyltransferase 2A gene (*KMT2A*) within chromosome band 11q23 has been reported in up to 10% of acute myeloid leukemia (AML) and myelodysplastic syndromes (MDS) and is associated with adverse risk [1,2]. *KMT2A*-PTD generates an elongated protein, usually by duplicating exons 2-8 or 2-10 of transcript NM_05933.4 (by convention referred to by their mutant junctions e8e2 or e10e2), however the underlying pathogenic mechanism is not well understood. *KMT2A*-PTD alone is insufficient to drive leukemogenesis and tends to occur after initiating mutations in the clonal hierarchy [3,4]. Although *KMT2A*-PTD has prognostic value, may predict response to targeted therapy, and could be a useful marker of measurable residual disease (MRD), it is not routinely incorporated into clinical practice due to the historic requirement of specialized RT-PCR assays for its detection [5,6,7].

Allelic complexity contributes to tumorigenesis through effects on oncogenes or tumor suppressors. In myeloid neoplasms, copy neutral loss of heterozygosity (CN-LOH) is a common mechanism of allelic complexity associated with adverse outcomes at several genomic loci including *FLT3, JAK2*, and *TP53*, while gains of structural variants have similarly shown clinical importance, such as duplications of *BCR-ABL1* associated with resistance in CML [8,9,10,11,12]. Few studies have directly investigated DNA allelic status of *KMT2A*-PTDs, given the traditional dependence on RNA-based assessment. Early work described *KMT2A*-PTD as affecting a single allele in the context of normal cytogenetics or trisomy 11, and a recessive gain-of-function effect was proposed through silencing of the wild-type allele [13,14,15]. A more recent study similarly predicted mono-allelic *KMT2A*-PTD as the typical state but also theorized the likelihood of occasional higher order gains, for which double duplication of exons or gain of a pre-existing *KMT2A*-PTD were proposed as mechanisms [3].

*KMT2A*-PTD detection has been integrated into next generation sequencing (NGS) panels, however allelic assessment remains minimally described [16,17,18,19]. Here, we develop approaches to integrate quantitative and allelic assessment of *KMT2A*-PTDs into standard targeted DNA NGS panel tests deployed in clinical and research settings. We explore multi-institutional data for evidence of *KMT2A*-PTD allelic complexity including CN-LOH and *KMT2A*-PTD gain, investigate emergence of allelic complexity in serial samples, and compare *KMT2A*-PTD burden to pathologic diagnosis and to RNA expression.

## Materials and Methods

### Sample Selection

DNA extracted from blood, bone marrow, or formalin-fixed paraffin-embedded tissue was tested by one of three targeted NGS panels: i) Heme SnapShot (HSS) based on anchored multiplex PCR (AMP; ArcherDx, Boulder, CO) performed clinically at Massachusetts General Hospital on unselected samples from 2017-2020 (MGH cohort: *n*=3700), ii) Rapid Heme Panel version 3 (RHP) based on NEBNext Direct capture/amplicon hybrid chemistry (NEB; New England BioLabs, Ipswich, MA) performed clinically at Brigham and Women’s Hospital on unselected samples from 2019-2020 (BWH cohort: *n*=5070), and iii) a myeloid-focused panel (MYP) based on hybrid-capture (HC) performed for research at Dana Farber Cancer Institute on a cohort of older AML patients (AML cohort: *n*=415), each followed by paired-end Illumina sequencing. Select clinical samples from the MGH cohort were concurrently tested by a targeted RNA-based NGS panel (Heme Fusion Assay: HFA) clinically validated to report pathogenic *KMT2A*-PTD isoforms (RNA/DNA cohort: *n*=350). The study was conducted in accordance with the Institutional Review Boards of each institution.

### Next generation sequencing and informatic analysis

Samples were processed by default pipelines of the DNA panels and underwent further customized informatics including batch-based copy number analysis (BR-CNV), SNP analysis, split-read analysis, and RNA-sequencing analysis. Details are provided in supplementary materials.

### Complex versus simple *KMT2A*-PTDs

The term complex *KMT2A*-PTD was used to refer to LOH (copy neutral or wild-type deletion) or multiple *KMT2A*-PTD copies in a single tumor cell. The relative copy number ratio of duplicated *KMT2A* exons over non-duplicated exons was 2 for LOH and between 1.5 and 2 for gains with more *KMT2A*-PTD copies than wild-type. The term simple referred to a single *KMT2A*-PTD copy without LOH, yielding ratios of 1.5 for diploid *KMT2A* and less than 1.5 for overall *KMT2A* gain. Assuming perfect accuracy, ratios above 1.5 in bulk NGS data thus implied at least a subclonal complex *KMT2A*-PTD component. Ratios above 2 implied a different mechanism and reflected higher order gains of the involved exons.

## Results

### Complex *KMT2A*-PTD events that increase the relative abundance of *KMT2A*-PTD include 11q23 gain of the mutant allele, broad CN-LOH, and focal CN-LOH

In order to identify *KMT2A*-PTD using targeted DNA NGS in unpaired tumor sequencing without a panel of normals, we introduced a batch-based ratio method (BR-CNV) for copy number analysis, where batch medians served as normal proxies given the relative infrequency of copy number changes across general hematologic samples. We validated the overall quantitative accuracy of BR-CNV against clinical FISH data and constitutional changes across various genomic loci (Supp Fig 1). For *KMT2A*-PTD exons, where orthogonal quantitative data from FISH was unavailable due to its cytogenetically cryptic nature, estimates using BR-CNV were similar across two DNA NGS assays with different library enrichment platforms and consistent with a clinical CNV pipeline (Supp Fig 2).

BR-CNV analysis revealed concurrent regional gain of 11q23 that originated from the *KMT2A*-PTD allele and not from the wild-type allele in 4% (4/94) of patients (P1-P4) with *KMT2A*-PTD (Supp Fig 3). One prototypical case (Figure 1A) exhibited copy number levels consistent with 5 copies of *KMT2A* exons 2-11 in ∼92% cells versus 3 copies of other *KMT2A* exons and 11q23 targets in ∼92% cells, thus implying 2 instances (duplication) of a *KMT2A*-PTD (2×2=4 copies of exons 2-11 and 2×1=2 copies of other regional targets) and 1 wild-type *KMT2A* allele (1 copy of all targets to reach the total 5 and 3 copies) per mutant cell. The regional *KMT2A* aberration was also detected by karyotype [dup(11)(q13q25)] and quantified at a similar level by *KMT2A* FISH (3 copies in 87% cells). In another case, we identified 2 discrete regional gain events affecting 11q23 with both originating from the *KMT2A*-PTD allele in ∼70% cells, thus yielding 3 instances of *KMT2A*-PTD and 1 wild-type *KMT2A* allele per mutant cell (Figure 1B). Its stepwise copy number profile suggested breakage-fusion-bridge cycles, consistent with its *TP53* mutation and globally complex karyotype.

**Figure 1.**
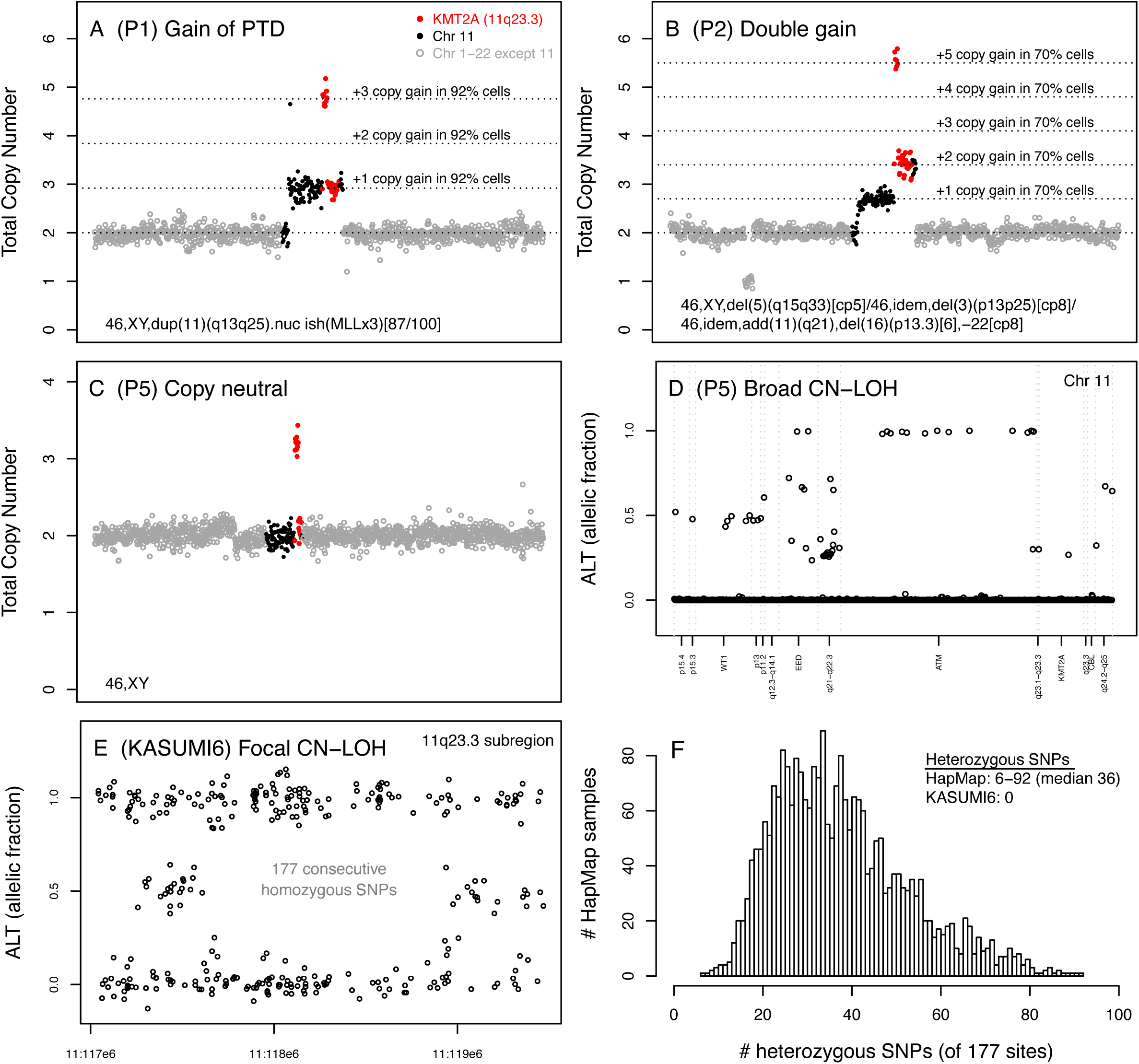
Patterns of *KMT2A*-PTD complexity from targeted DNA NGS data. (A) Gain of 11q23.3 from the *KMT2A*-PTD allele: P1 had bulk copy number levels consistent with 5 copies of *KMT2A* exons 2-11 in ∼92% cells and 3 copies of distal *KMT2A* exons in ∼92% cells, indicating 11q23.3 gain from the *KMT2A-PTD* allele (as opposed to gain from the wild-type allele, which would correspond to 4 copies of exons 2-11 and 3 copies of distal *KMT2A*). (B) Multiple gains of 11q23.3 from the *KMT2A*-PTD allele: P2 had a *TP53* mutation, complex karyotype, and step-wise copy number profile over 11q, consistent with a *KMT2A*-PTD of exons 2-8 subject to sequential distal gain events spanning at least targets of (i) 11q22.2-q23.3 followed by (ii) 11q23.3, occurring in ∼70% of cells based on copy number levels. (C-D) Broad CN-LOH affecting 11q: P5 had a normal karyotype, a flat copy number profile across chromosome 11 by BR-CNV, and allelic imbalance spanning targets of 11q by SNP analysis; the integrated data thus indicated 11q CN-LOH, in contrast to prior studies reporting solely mono-allelic involvement in cytogenetically normal cases. (E-F) Focal CN-LOH affecting *KMT2A*: KASUMI6 demonstrated 0 heterozygous SNPs over 177 consecutive sites of the CytoscanHD array between 11:117,619,027-118,938,315 (∼1.3 Mb) containing *KMT2A* but not *CBL*, whereas 2500+ HapMap genomes had a median of 36 heterozygous SNPs and never 0 (range 6-92) over these 177 sites. Combined with the high copy number gain of exons 2-8 beyond the level of a simple *KMT2A*-PTD, the findings argued in favor of interstitial CN-LOH. RNA expression was also accordingly increased compared to the simple *KMT2A*-PTD cell line EOL1 (Supp Fig 8).

**Figure 2.**
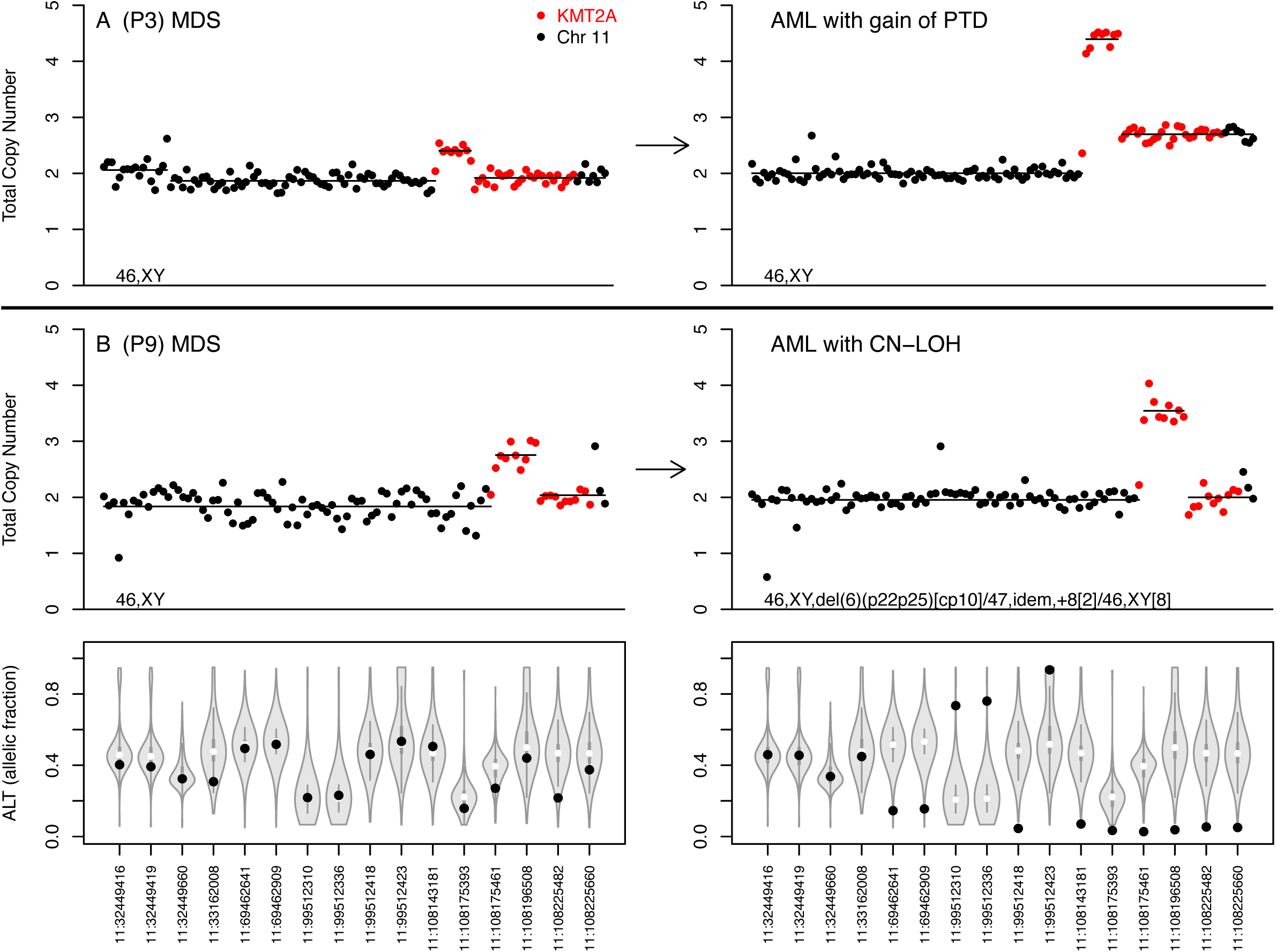
Emergence of *KMT2A*-PTD complexity during progression to AML. (A) In P3, 11q23.3 gain complexity emerged as the predominant clone at a late time point in a rapidly progressing AML that evolved from MDS with *KMT2A*-PTD (see also Figure 3). Complexity was not detected at the intermediate time point of AML diagnosis (not shown here; see Supp Fig 3C), however its subclonal presence below the limit of bulk NGS detection could be inferred at AML diagnosis from predicted clonal hierarchy relative to an *NRAS* hotspot variant (Supp Fig 6A). (B) In P9, CN-LOH complexity emerged in a *KMT2A*-PTD during AML progression from MDS. Integrated BR-CNV and SNP analysis at the AML time point showed a flat copy number profile with allelic imbalance of multiple SNPs across 11q centromeric to *KMT2A*, where CN-LOH was inferred to extend to *KMT2A* given the high amount of copy number gain beyond that of a simple *KMT2A*-PTD. Allelic imbalance of SNPs was detected relative to empirical distributions given biases of the clinical NGS assays. Occasional clinical cases lacked informative SNPs and were equivocal due to the more limited coverage of the clinical assays. We thus remark that quantifying the level of allelic imbalance in the setting of these biases, intrinsic noise, and sparsity of informative SNPs was challenging and not performed (e.g. to estimate subpopulations).

We next evaluated for broad CN-LOH within 11q, defined as affecting the whole arm or a large telomeric segment, as another potential mechanism of increased relative abundance of *KMT2A*-PTD. By integrating analysis of chromosome 11 SNPs, we found CN-LOH in 11 of 94 (12%) patients with *KMT2A*-PTD (P5-P15) (Figure 1C-D, Supp Figs 4-5). *KMT2A*-PTD accounted for 36% of all 11q CN-LOH cases from our AML cohort and were mutually exclusive with 11q CN-LOH that harbored *CBL* mutations (45% of all 11q CN-LOH), indicating that 11q CN-LOH in AML is largely explained by mutually exclusive alterations in *KMT2A*-PTD and *CBL*.

In some cases, we noted high *KMT2A*-PTD copy number levels indicative of complexity but without evidence of broad CN-LOH or 11q gain. To determine whether small interstitial CN-LOH beyond the resolution of our targeted NGS panels might conceivably underlie such cases, we analyzed publicly available genomic cell line data. The KASUMI6 AML cell line has been noted to exhibit gain of *KMT2A* exons 2-8 at a high normalized log2 ratio (+0.9) based on Affymetrix SNP 6.0 array data from the Cancer Cell Line Encyclopedia (CCLE) [3]. We used Rawcopy to re-process recent public raw SNP data of KASUMI6 generated on the denser Cytoscan HD array [20, 21]. Broad CN-LOH was not seen within 11q, however Rawcopy predicted a stretch of 177 consecutive homozygous SNPs on the array across ∼1.3 Mbp from 11q23.3 containing the ∼90 Kbp locus of *KMT2A* and not *CBL* (Figure 1E). By comparison, no samples among 2584 HapMap genotypes were homozygous over all 177 SNP sites, where heterozygous SNPs ranged between 6-92 for a median of 36 per genotype, thus supporting interstitial CN-LOH (Figure 1F). Given the availability of concurrent RNA-seq data from CCLE (SRX5414390 / SRR8615363), we also verified the presence of aberrant RNA reads spanning the *KMT2A* exon8:exon2 mutant splice junction to exclude the rare possibility of non-PTD rearrangements represented by gain of *KMT2A* exons 2-8 [22,23]. These mutant reads were uncharacterized and not aligned to the *KMT2A* locus in the processed bam files but were present in the raw FASTQ files at a similar level to reads spanning exon8:exon9 or exon1:exon2 wild-type junctions and moreover persisted after polyA enrichment in the RNA-seq protocol, thus consistent with transcripts from genomic duplication (*KMT2A*-PTD) versus the alternative possibility of backspliced circular RNAs.

### *KMT2A*-PTD complexity is acquired as a subclonal event at progression or relapse

To determine when *KMT2A*-PTD complexity develops in clonal evolution, we analyzed serial samples from 7 patients with complexity (P3-P4, P9-P13). In 5 of 7 patients, complex *KMT2A*-PTD events emerged from previously documented simple *KMT2A*-PTD (4 patients) or wild-type *KMT2A* (1 patient) near the time of secondary AML transformation or AML relapse, characterized by both 11q23 gain (P3-P4, Supp Fig 3C-D) and broad 11q CN-LOH (P9, P11-P12, Supp Fig 5A-C). To quantify the relative proportion of simple and complex clones, we decomposed serial samples with 11q23 gain into estimated wild-type, simple, and complex subpopulations to match bulk copy number levels (Fig 3, Supp Fig 3E). In one patient (P4), the simple component diminished over time as the complex component expanded, indicating that a subclone harboring a complex *KMT2A*-PTD had a competitive advantage over the parental clone with simple *KMT2A*-PTD. In the second patient (P3), the bone marrow evolved relatively rapidly from a mixed simple/wild-type state to a mixed complex/simple state, however further time points were not tested to characterize subsequent evolution of the complex component. Analogous decomposition of the CN-LOH cases into subpopulations was not possible due to assay biases. The 7 patients were associated with 3 deaths due to AML occurring 51-120 days after detection of complexity and 221-434 days after diagnosis, 1 death due to GVHD, and 3 cases of stable remission. Three more complex *KMT2A*-PTD cases without serial samples had limited clinical follow-up, revealing 2 more deaths due to AML and 1 more case of stable remission.

**Figure 3.**
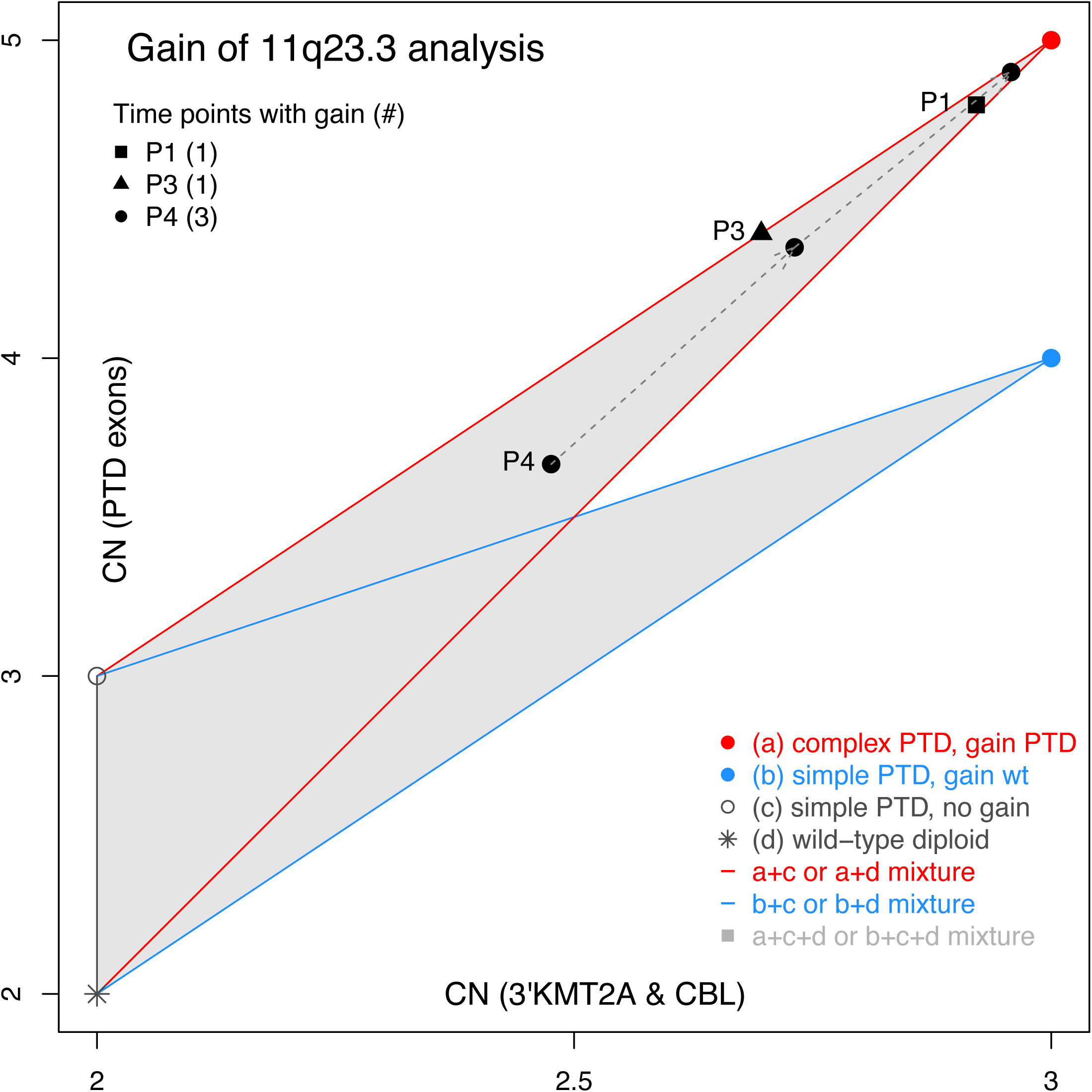
Gain of 11q23.3 analysis. By definition, a single *KMT2A*-PTD cell with 11q23.3 gain has 3 copies of distal *KMT2A* (representing 11q23.3) and is either complex (red circle) or simple (blue circle) depending on whether gain originates from the *KMT2A*-PTD or wild-type allele, yielding 5 or 4 copies of PTD exons respectively. By contrast, a standard simple *KMT2A*-PTD cell (black open circle) without 11q23.3 gain has 2 copies of distal *KMT2A* and 3 copies of PTD exons, and a wild-type diploid cell (black asterisk) has 2 copies of each. Since copy number behaves linearly, mixed populations containing 2 components exist on the line connecting those components, while mixed populations containing 3 components exist within the corresponding triangle (i.e., more generally, mixed populations are represented by the convex hull of their components). P1, P3, and P4 all resided in regions of the plot indicative of a major complex *KMT2A*-PTD component characterized by gain from the *KMT2A*-PTD allele and minor components of standard simple *KMT2A*-PTD (without gain) and wild-type. P3 existed essentially on the upper line and corresponded to approximately 70% complex *KMT2A*-PTD and 30% standard simple *KMT2A*-PTD. P4 started at approximately 48% complex *KMT2A*-PTD, 23% standard simple KMT2A-PTD, and 29% wild-type, and the complex component grew over time reaching ∼96% at the final time point. P2 (off the plot) is shown in Supp Fig 3 along with more details and earlier non-complex time points of P3 and P4.

We observed that *KMT2A*-PTD complexity was often associated with blast proportion over the course of disease progression, sometimes as the sole molecular change (P3, Supp Fig 6A) and sometimes with concurrent gene mutations. When *KMT2A*-PTD complexity emerged alongside other gene mutations, we inferred clonal hierarchy by estimating the fraction of clonal cells involved by *KMT2A*-PTD complexity relative to other mutations. In P3, this enabled the inference of subclonal *KMT2A*-PTD complexity at time of AML diagnosis below the limit of our CNV-based limit of detection, followed by rapid expansion (Supp Fig 6A). In two other exemplar cases (P4 and P6, Supp Fig 6B-C), we determined that *KMT2A*-PTD gain or CN-LOH was subclonal to other biallelic gene mutations (*NF1, RUNX1*), indicating that acquisition of KMT2A-PTD complexity may drive terminal transformation.

### *KMT2A*-PTDs with high copy number indicative of complexity are enriched in AML over MDS and associated with greater RNA expression

Copy number ratios of *KMT2A*-PTD exons relative to distal *KMT2A* were higher in AML (mean 1.64, n=73) compared with MDS (mean 1.40, n=25) (p=0.00006 by t-test) (Figure 4). Ratios in AML clustered in 2 main groups predicted to correspond roughly to simple and complex *KMT2A*-PTDs, and the dip test for unimodality supported a unimodal distribution for MDS (p=0.843) and a multimodal distribution for AML (p=0.027) [24]. We compared the proportion of high versus low copy ratio *KMT2A*-PTD in MDS versus AML, using an empirically defined cutoff of 1.6, which corresponded to a natural theoretical threshold relative to the discrete ratios of 100% clonal *KMT2A*-PTDs: 1.5 (simple diploid) versus 1.67 (complex: *KMT2A*-PTD duplication) and 2 (complex: CN-LOH). We found that high copy ratios consistent with complexity were significantly more common in AML (33 of 73; 45%) compared with MDS (3 of 25; 12%) (p=0.003; Fisher’s exact test). Of 3 high ratio MDS cases, 2 had complexity present at the initial time point and subsequently progressed to AML (P10, P15), suggesting a potential prognostic utility of high ratios in MDS.

**Figure 4.**
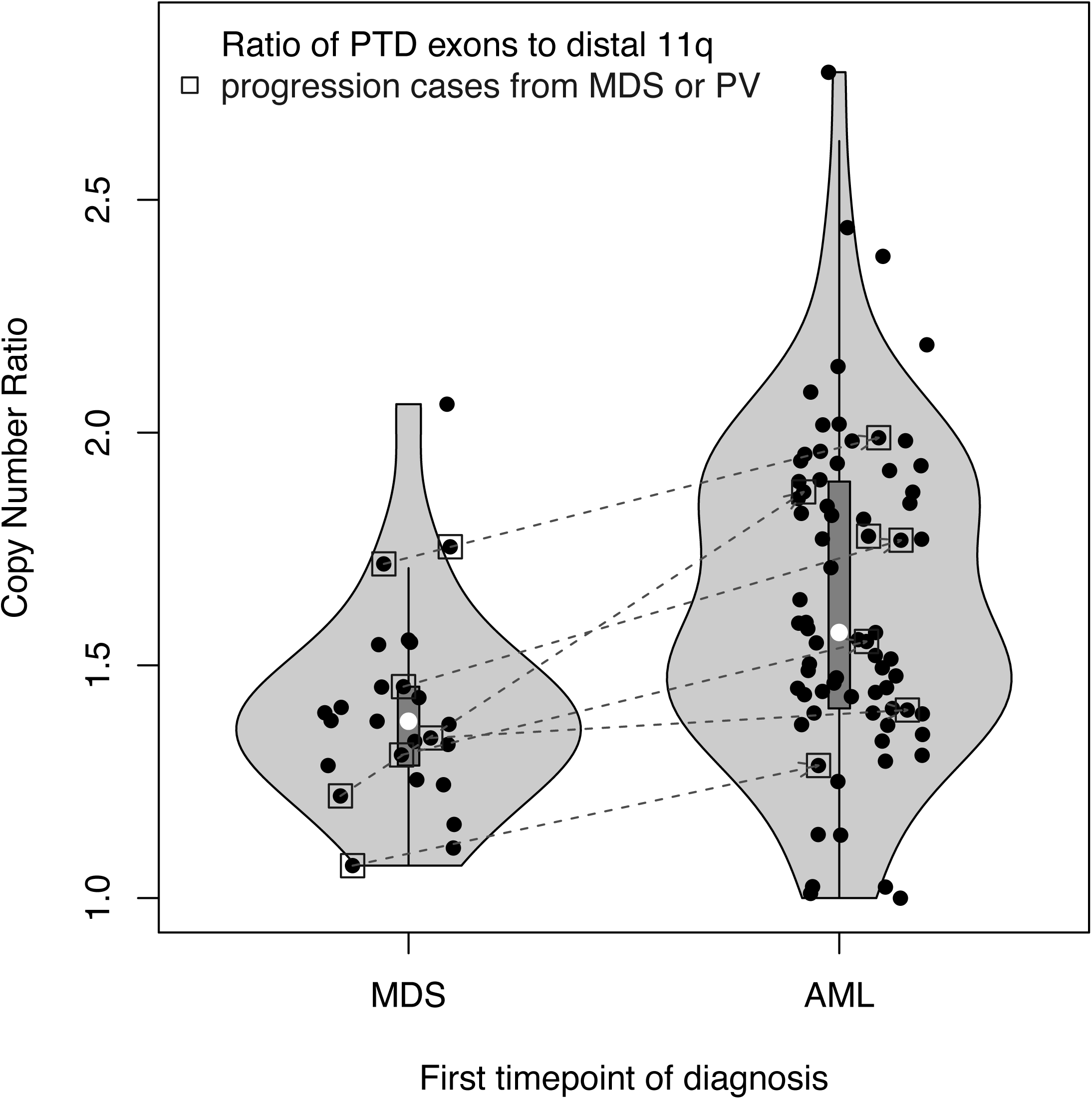
Copy number ratios of *KMT2A*-PTDs at first available time points by diagnosis. Ratios were calculated from either BR-CNV segmentation levels of *KMT2A*-PTD exons over adjacent distal 11q targets or split-read estimates in cases without a BR-CNV signal. The *KMT2A*-PTD cohorts contained *N*=94 patients with diagnoses of AML (73), MDS (25), and unknown (2), where 6 patients progressed from MDS to AML while 1 patient progressed from polycythemia vera (PV) to AML with its *KMT2A*-PTD emerging only at the AML time point. Median and mean ratios were 1.57 and 1.64 in AML and 1.38 and 1.40 in MDS. Squares: cases with secondary AML transformation from MDS or PV. Box plots within violin plots: white dots represent medians, rectangles correspond to interquartile ranges, and whiskers have length equal to 1.5 times interquartile range or end at most extreme outliers.

By combining with results of the previous section, we examined prevalence and characteristics of *KMT2A*-PTD complexity among *KMT2A*-PTD positive MDS cases experiencing secondary AML transformation, acknowledging limited numbers in our cohort. The majority of such cases (5/7; 71%) had complexity either at diagnosis (P10, P15) or near progression (P3, P9, P11), and were associated with e8e2 or e10e2 isoforms. The remaining minority (2/7; 29%) had copy ratios and clone sizes consistent with persistent simple *KMT2A*-PTDs and were characterized by e6e3 or e13e3 isoforms. High copy ratios were more prevalent in e8e2 than e10e2 and other *KMT2A*-PTDs, raising the possibility of biological variability across different isoforms (Supp Fig 7).

*KMT2A*-PTD complexity, resulting in high copy ratio, may exert its effect by increasing the relative expression of the mutated allele. Therefore, we investigated the relationship between RNA expression levels and DNA-based allelic burden of *KMT2A*-PTDs. We compared KASUMI6, harboring a complex *KMT2A*-PTD (CN-LOH) involving exons 2-8 with an estimated log2 ratio of +0.9, to the well-studied EOL1, which similarly harbored a *KMT2A*-PTD involving exons 2-8 but with an estimated log2 ratio of +0.4 consistent with simple status [3]. RNA-seq data from KASUMI6 accordingly had more reads spanning the exon8:exon2 mutant junction than EOL1 (136 versus 37 reads) and a greater ratio relative to reads spanning the exon8:exon9 wild-type junction (0.70 versus 0.24); similarly, ratios of *KMT2A*-PTD mutant junctions to wild-type junctions in clinical targeted RNA-seq data generally correlated with DNA-based allelic burden (Supp Fig 8).

### *KMT2A*-PTDs are specific to MDS and AML in an unselected cohort of hematologic conditions and are effectively detected and quantified by NGS panels

We next evaluated the frequency and characteristics of *KMT2A*-PTD across an unselected, sequential cohort of 476 uniformly sequenced patients with hematologic disease. Consistent with prior studies [3], *KMT2A*-PTDs were present only in AML (10 of 165, 6.1%) and MDS (5 of 49, 10.0%), but not in other myeloid diseases (Ph-negative myeloproliferative neoplasms, CML, or CNL), lymphoid diseases (acute lymphoblastic leukemia, non-Hodgkin lymphoma, myeloma, hairy cell leukemia, LGL leukemia) or non-clonal hematologic diseases (aplastic anemia, HLH) (Supp Fig 9). We compared the performance of DNA-based NGS detection of *KMT2A*-PTD relative to available clinical RNA results. In the RNA/DNA cohort, BR-CNV had a sensitivity of 90% (18/20) and a specificity of 100% (330/330) relative to clinically validated RNA-based testing (HFA) for detection of pathogenic *KMT2A*-PTD isoforms; combining with split-read analysis resulted in 95% (19/20) sensitivity and 100% (330/330) specificity, where the lone false negative was associated with a low allelic fraction inferred from size of co-mutations. Split-reads provided another method to estimate allelic burden when available. Since traditional validation of split-read based accuracy was not possible due to lack of an orthogonal quantitative clinical assay, a limited proof of principle was instead established through (non-PTD) *KMT2A* rearrangements quantified by clinical FISH testing (Supp Fig 10A). Split-read based estimates of *KMT2A*-PTD allelic burden moreover correlated well with BR-CNV estimates (Supp Fig 10B).

### Split-read analysis reveals novel and atypical *KMT2A*-PTDs and potentially enables patient-specific MRD evaluation

Genomic breakpoints of *KMT2A*-PTDs were occasionally sequenced by the NGS panels and aligned as split-reads, depending on their proximity to targeted *KMT2A* exons. Split-reads captured 25 unique breakpoint-pairs from 26 patients, consistent with both well-known pathogenic *KMT2A*-PTD isoforms and potential novel isoforms (e13e3, e15e2, e6e2) (Figure 5). The e6e2 isoform was atypical for being out-of-frame although theoretically might generate in-frame e6e3 transcripts from alternative splicing. The cohorts demonstrated the predicted RNA isoforms e8e2 (n=43), e10e2 (34), e8e4 (4), e9e2 (4), e11e2 (3), e6e3 (2), e13e3 (2), e6e2 (1), and e15e2 (1) based on split-reads and BR-CNV. Split-read based estimates were occasionally helpful in *KMT2A*-PTD evaluation and interpretation; the novel e15e2 case (AML) was negative by BR-CNV, however split-reads enabled allelic fraction estimates of 1.0-3.0% across serial samples, consistent with a minor *KMT2A*-PTD subclone relative to moderate blast percentages and clonal mutations in *TP53* and *DDX41* (Supp Fig 11A). Clinical significance was thus uncertain, given its different behavior compared to typical pathogenic *KMT2A*-PTDs that recurrently arise in dominant clones as early cooperating mutations critical to AML development [3].

**Figure 5.**
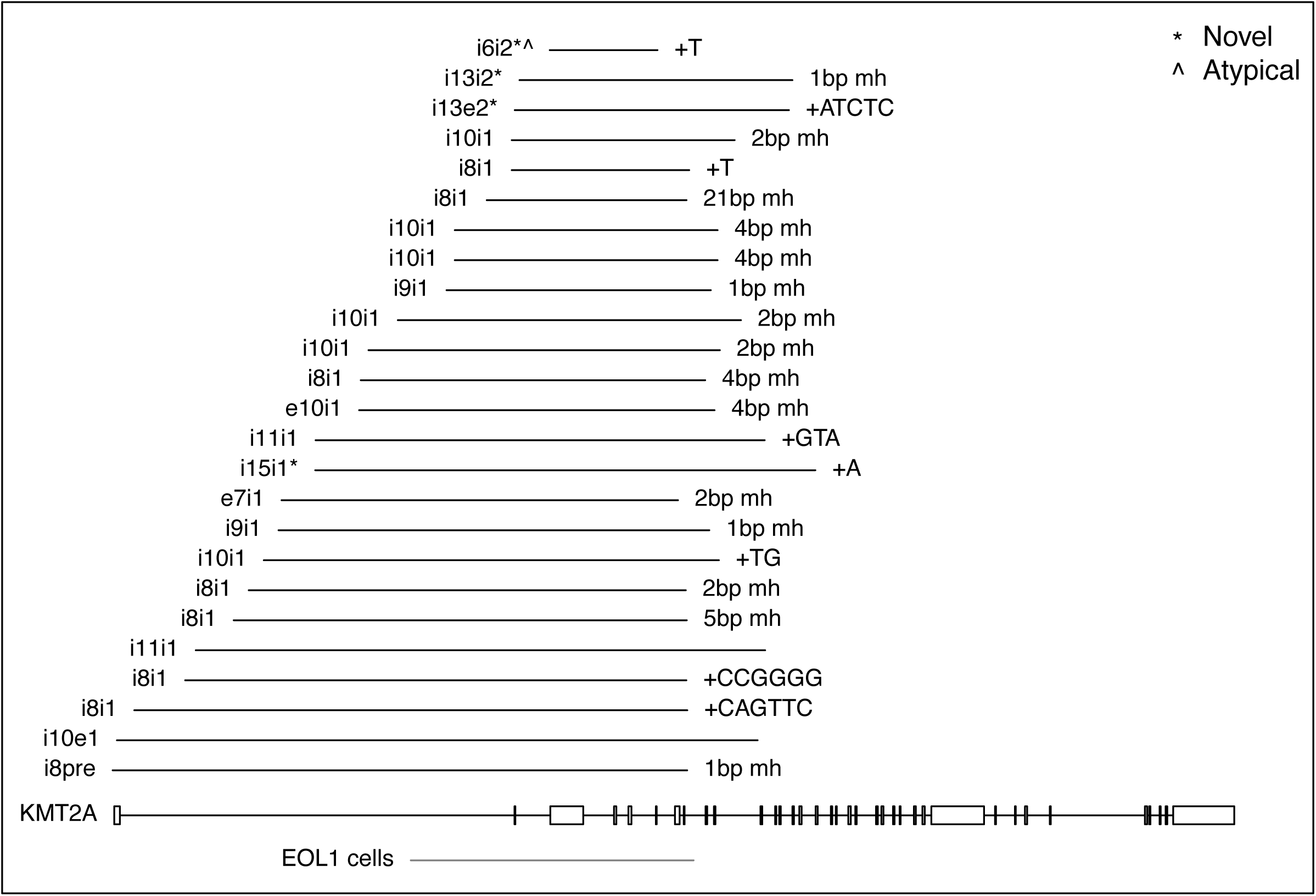
*KMT2A*-PTD genomic breakpoints detected by split-reads. Novel isoforms e13e3 (2 patients), e15e2 (1 patient), and e6e2 (1 patient) were predicted from breakpoints in i13e2 and i13i2, i15i1, and i6i1 respectively. The isoform e6e2 would be out-of-frame and thus atypical, raising the question of whether e6e3 transcripts might be generated by alternative splicing. The breakpoint i8pre was predicted to generate the e8e2 isoform given the lack of a 3’ splice acceptor site before exon 1. +XXX: intervening exogenous filler sequence. mh: microhomology. e: exon. i: intron. pre: 5’ of *KMT2A*.

Serial samples experiencing relapse in our cohorts demonstrated *KMT2A*-PTD persistence, consistent with prior studies proposing *KMT2A*-PTDs as stable markers of MRD detection. In contrast to the limited sensitivity of copy number methods under low tumor cellularity, split-reads showed promise for patient-specific MRD assessment in select cases, including instances of partially intraexonic breakpoints within targeted regions that yielded considerable sequencing depths. Samples from 10 patients were negative for *KMT2A*-PTDs by BR-CNV but positive by split-reads, all occurring in the context of lower blast percentages except for the subclonal e15e2 case described earlier; four were associated with earlier or later cellular samples positive by BR-CNV. The presence of a split-read regardless of coverage depth remained specific, especially in the context of a known breakpoint, although quantitative accuracy might be compromised. A second case of *DDX41*-related AML, with *KMT2A*-PTD at diagnosis by BR-CNV but only rare i10i1 split-reads due to shallow coverage, was subsequently positive for MRD well below the resolution of BR-CNV (5.2% blasts with *TP53* and *DDX41* variants at 1.7% and 0.8%); although the i10i1 breakpoint was covered by only 10 reads at the MRD time point, one was a split-read of the previously characterized mutant junction (Supp Fig 11B) consistent with its persistence.

An unusual breakpoint occurred within *KMT2A* intron8/pre-exon1 and corresponded to BR-CNV gain of exons 1-8; it was predicted to generate e8e2 transcripts given the lack of a splice acceptor in exon 1 (Supp Fig 12A-B). Without the help of split reads, its copy number pattern of partial duplication of the 5’ segment of *KMT2A* would have been difficult to resolve from *KMT2A* rearrangement with partial 5’ gain, which is recurrent in AML and occurred in our cohorts (Supp Fig 12C-D) [25,26,27]. It is worth recognizing other pitfalls of copy number methods to detect *KMT2A*-PTD, where split-reads may also facilitate characterization (Supp Fig E-H).

## Discussion

Allelic state can have important clinical implications in myeloid neoplasms, as epitomized by *FLT3*-ITDs, whose quantification by allelic ratio is part of European LeukemiaNet risk stratification and reflects both allelic state and clonal burden [28]. Direct 13q LOH detection is also effective at identifying high-risk *FLT3*-ITDs, and homozygosity at relapse has been linked to unrecognized subclonal populations at diagnosis [10,29,30]. Along similar lines, we used targeted DNA NGS to characterize *KMT2A*-PTD allelic state and estimate allelic burden, revealing the explicit emergence and outgrowth of *KMT2A*-PTD complexity in multiple cases of secondary transformation to AML and an overall enrichment of inferred complexity in AML compared to MDS. Subtle suggestions of *KMT2A*-PTD complexity have also arisen in the literature. *CBL* mutations were previously reported in 58% (7/12) of general myeloid cases with 11q CN-LOH whereas homozygous *JAK2* V617F, *FLT3*-ITDs, and *RUNX1* mutations have been identified in nearly all cases of CN-LOH affecting 9p, 13q, and 21q respectively [30,31,32,33]. We accordingly found mutually exclusive *KMT2A*-PTD and *CBL* variants in 36% (4/11) and 45% (5/11) of AML cases with broad CN-LOH affecting 11q, indicating that *KMT2A*-PTD underlies a portion of unexplained 11q CN-LOH. In 2 prior studies, unexplained 11q CN-LOH without *CBL* mutations was reported in 1% of MDS (1/95 and 1/108) versus 2-4% of AML (3/143 and 1/28), suggesting soft support for our finding of enriched inferred complexity (CN-LOH) in AML over MDS (assuming relatively equal *KMT2A*-PTD prevalence in MDS and AML and that unexplained cases predominantly harbor *KMT2A*-PTD) [31,34].

Although CCLE contained only 2 *KMT2A*-PTD positive AML cell lines, we established evidence of complexity in 50% (1/2; KASUMI6). Similarly, we inferred allelic complexity in 45% (33/73) of *KMT2A*-PTD positive AML at diagnosis via high copy ratios, despite explicit characterizations in only 16% (15/94) of patients with *KMT2A*-PTD. Technical limitations may underlie this discrepancy between inferred and explicit complexity, including (1) focal CN-LOH beyond the resolution of NGS panels as found in KASUMI6, (2) underestimation of broad CN-LOH due to limited genomic targets resulting in indeterminate cases lacking informative heterozygous SNPs, and (3) the possibility of uncharacterized mechanisms of complexity that may underlie unusually high copy ratios. Given our substantial prevalence estimates, a few reasons may explain why *KMT2A*-PTD allelic complexity has received sparse attention in the literature. First, assessment of *KMT2A*-PTD has historically used assays based on RNA and not DNA. Second, early studies described *KMT2A*-PTD as affecting a single allele and found evidence of epigenetic silencing of wild-type alleles to explain RNA-expressed allelic imbalance [13,14].

Since *KMT2A*-PTD is insufficient to drive leukemogenesis by itself, allelic complexity may be one of many factors promoting leukemogenesis within *KMT2A*-PTD positive MDS, potentially through dosage effects; we thereby confirmed RNA levels to be correlated with allelic burden. A recent study linked high initial *KMT2A*-PTD RNA levels to poor disease outcomes, however other studies have reported conflicting results [5,35,36]. KASUMI6, derived from a relapsed AML with a dominant negative *CEBPA* mutation, may provide a useful model to better understand pathogenicity of *KMT2A*-PTD complexity, whereas many *KMT2A*-PTD studies have relied on EOL1, which harbors a simple *KMT2A*-PTD and was derived from a chronic eosinophilic leukemia with a *FIP1L1-PDGFRA* fusion [6,37].

Risk stratification by genetic subgroups of *KMT2A*-PTD positive MDS or AML has been the subject of several studies, however assessment of co-mutational associations of *KMT2A*-PTD complexity was beyond the scope of this study. Interesting behaviors related to *IDH2* were nevertheless observed. All 5 *KMT2A*-PTD positive MDS cases harboring *IDH2* mutations progressed to AML, with 4 demonstrating *KMT2A*-PTD complexity, whereas prior studies of *IDH2*-mutant MDS described a progression rate of 23% (10/43) [38]. Given the heavy biases of our cohorts, larger unbiased studies would be beneficial. Our cohorts included an *IDH2*-mutant polycythemia vera (PV) with emergence of *KMT2A*-PTD and CN-LOH upon AML progression. *IDH2* mutations were previously reported in 1.4% (6/421) PV and 8.3% (1/12) post-PV AML, indicating greater prevalence in blast-phase disease [39]. Our example suggests *KMT2A*-PTD complexity as a candidate driver. The 2 highest *KMT2A*-PTD burdens in our cohorts also had *IDH2* mutations and CN-LOH. The highest ratio (2.77) was entirely attributable to a single mutant junction based on *KMT2A*-PTD split-read estimates, raising the possibility of episomal amplification [40,41]. Specialized FISH probes may be capable of providing supporting evidence. Co-mutational frequencies in our *KMT2A*-PTD cohort were generally similar to prior studies, however we described 2 *DDX41*-related AML cases with *KMT2A*-PTD, minimally found in the literature

We provided approaches to integrate quantitative and allelic assessment of *KMT2A*-PTDs into targeted DNA NGS. Copy-number and SNP based assessment was generally effective at detection and relatively effective at allelic characterization in diagnostic samples but became insensitive at low tumor cellularity, whereas a split-read approach yielded lower limits of detection when coverage was adequate and potentially facilitated patient-specific MRD testing but was insufficient to characterize complexity. Enhancing split-read detection through targeting of *KMT2A* intronic breakpoint regions may improve sensitivity and specificity [42]. Split-reads also enabled characterization of novel isoforms, including a rare e15e2 case with the unusual property of being a minor subclone. Larger datasets will be required to resolve the possibility of differential behavior or propensities for complexity by isoform. Early identification of complex *KMT2A*-PTD subclones may ultimately prove useful for clinical decision-making, given our examples of emergent *KMT2A*-PTD complexity during progression and relatively rapid outgrowth of subclones harboring complexity. However, exact time points of emergence could not be determined by our current bulk NGS methods, thus the ability to recognize low-level allelic complexity remains a significant challenge requiring single cell or other new techniques.

## Data Availability

Select data produced in the present study are available upon reasonable request to the authors.

## Acknowledgements

This work was supported by the National Institutes of Health [K08CA204734 (RCL), K08CA263555 (CJG), and P30CA016056 (EJW; involving the use of Roswell Park Comprehensive Cancer Center’s Hematologic Procurement Shared Resource)], the American Society of Hematology (HMM), the Ted and Eileen Pasquarello Tissue Bank in Hematologic Malignancies, the Dana-Farber Cancer Institute Center for Cancer Genome Discovery, and the Dana-Farber Cancer Institute Hematologic Malignancy Data Repository (Anne Charles, Kevin Copson, Nish Patel).

## Author Contributions

HKT and RCL designed the study and wrote the manuscript. CJG, HMM, MHH, ESW, LPG, ASK, VN, and RCL contributed data. HKT, CJG, and PD developed informatic tools. All authors performed data analysis and edited the manuscript.

