## Supplementary Material for "Allelic Complexity of *KMT2A* Partial Tandem Duplications in Acute Myeloid Leukemia and Myelodysplastic Syndromes"

**Corresponding Author:**

R. Coleman Lindsley, MD, PhD

**Supplementary Methods**

DNA sequencing of *KMT2A* and targeted genes: The NGS panels targeted regions of 95-114 genes (HSS v2-v4), 88 genes (RHP v3), or 117 genes and 215 intergenic/SNP loci (MYP), including *KMT2A* exons 1-36 (HSS) or exons 1-13, 24-26, and 28-30 (RHP, MYP) relative to transcript NM_005933.4. Raw sequencing results were processed by clinical or research informatic pipelines consisting of alignment to the hg19 human genome reference through Novoalign v3.0.7.00 (HSS), bwa mem v0.7.17 (RHP), or bwa v0.5.9 (MYH), deduplication by Picard MarkDuplicates v1.128 (HSS) or v1.130 (MYH), reduction of 8 bp unique molecular indexes via fgbio v0.4.0 (RHP), and SNV/indel detection by an ensemble approach (HSS; MuTect v1.1.7, LoFreq v2.1.2, GATK vnightly-2016-01-24-gaa090b7, laboratory developed hotspot caller), VarDict v1.6.0 (RHP), or VarScan v2.3.3 (MYH), where the clinical assays were validated to detect SNV/indels at allelic frequencies of 10% (HSS) or 3% (RHP). *FLT3*-ITD detection was performed by FLT3_ITD_ext v1.1 (HSS, RHP, MYH) and clinically validated for RHP [1]. Copy number variation was assessed and clinically validated through an internally developed algorithm RobustCNV for one of the assays (RHP) and was not performed in the standard pipelines of the other assays (HSS, MYH) [2,3]. Processed results from the informatic pipelines were reviewed manually by molecular pathologists for clinical reporting (HSS: SNV/indels, RHP: SNV/indels and CNV) or underwent internal evaluation (MYH).

Batch Ratios CNV method (BR-CNV): Of the 3 assays, RHP was the only one with a clinically validated pipeline for CNV evaluation, based on a panel of normals (PON), while HSS and MYH did not have CNV assessment or associated sequenced PONs. We thus applied an alternative approach across all three assays. Relative copy numbers were assessed through a batch-based method BR-CNV applied to sequencing batches of 5-27 (HSS), 31 (RHP), and 88 (MYH) samples and predicated on 2 main underlying assumptions: (1) linearity of sequencing read depths for a given target locus relative to copy number (or more specifically, input DNA) and (2) a diploid state over each target locus in the majority of samples of a sequencing batch. Assumption (1) could be evaluated per locus with removal of poorly performing targets, while assumption (2) could only be checked on a broad chromosomal level through concurrent karyotypes and had to be accepted at face value for each specific target, relying instead on the tendency for hematologic genomes to be relatively stable.

For each sample $S$ of a batch and targeted region $R$ specified in assay design bedfiles, mean coverage depth $D_S(R)$ was generated by the clinical and research informatic pipelines from alignments of raw reads (HSS), UMI-consensus reads (RHP), or deduplicated reads (MYH), where counts were extracted from alignment files by either samtools (HSS, RHP) or by Picard CollectHsMetrics (MYH). Each pair of regions $R_j$ and $R_k$ was then associated with a coverage ratio $X_{jkS} = D_S(R_j) / D_S(R_k)$ and a median-adjusted coverage ratio $Y_{jkS} = X_{jkS} / median_T\{X_{jkT}\}$ relative to samples $\{T\}$ in the batch. The relative copy number based on batch ratios of targeted region $R_a$ in sample $S$ was defined as

$$\text{BR-CNV}_S\,(R_a) = median_k \{ Y_{akS} / median_j\{Y_{jkS}\} \}$$

For efficiency, it was also possible to fix the value of $k$, as long as it corresponded to a robust target region $R_k$, with minimal impact on relative copy numbers.

Relative copy numbers were log2-transformed and underwent circular binary segmentation (CBS) using the R Bioconductor package DNAcopy (Olshen 2004: 15475419). Both chromosome 11 and a smaller region around *KMT2A* were separately processed by CBS to have a higher sensitivity for focal copy number changes, and segmentation results with breakpoints in *KMT2A* were manually reviewed. Segmentations with focal gain within the 5' end of *KMT2A* in the absence of copy number loss of 3' *KMT2A* were considered strong evidence *KMT2A*-PTDs when the gain affected a subset of contiguous exons within 2 to 15, and were considered possible *KMT2A*-PTDs when the gain included exon 1. The *KMT2A*-PTD copy number burden was defined as the difference between the segmentation level of the gained segment minus either (i) the average of the surrounding segmentation levels (in most cases) or (ii) the segmentation level of 3' *KMT2A* in rare instances of definitive distal 11q gain.

BR-CNV was further modified to optionally integrate a panel of normals (PON) when available. RHP had an available PON which consisted of 2 sub-batches $PON_0$ and $PON_1$ that were sequenced during the original validation of the RHP assay The overall strategy was to adjust coverage ratios of the normal samples in $PON_0$ and $PON_1$ by scaling to match medians of any given clinical batch, and then to use unadjusted normals, adjusted normals, and the clinical batch median to determine a “nearest normal” to a clinical sample and finally calculate copy numbers relative to this nearest normal. Specifically, coverage ratios of normal samples in $PON_0$ and $PON_1$ were scaled respectively by the multiples $c_{jk}$ = ($median_T\{X_{jkT}\}/median_N\{X_{jkN}\}$) and $c'_{jk}$ = ($median_T\{X_{jkT}\}/median_{N'}\{X_{jkN'}\}$) where {$N$}, {$N'$}, and {$T$} denote all samples of $PON_0$, $PON_1$, and the clinical batch. Given a clinical sample S, the "nearest normal" (NN) was then defined in terms of the coefficients $\{p^*_N, p^*_{N'}, q^*_N, q^*_{N'}, r^*\}$ solving the linear program:

$$\text{Minimize} \sum_{j,k}\left(X_{jkS} - \left(\sum_{N} p_N X_{jkN} + \sum_{N'} p_{N'} X_{jkN'} + \sum_{N} q_N c_{jk} X_{jkN} + \sum_{N'} q_{N'} c'_{jk} X_{jkN'} + r\,[median_T\{X_{jkT}\}]\right)\right)^2$$

$$\text{where} \sum_{N} p_N + \sum_{N'} p_{N'} + \sum_{N} q_N + \sum_{N'} q_{N'} + r = 1 \text{ and all } p_N \geq 0,\ p_{N'} \geq 0,\ q_N \geq 0,\ q_{N'} \geq 0,\ r \geq 0.$$

The PON-based relative copy number is finally:

$$\text{PON-BR-CNV}_S\ (R_a) = median_k\ \{\ X_{akS}\ /\ NN_{akS}\ \}$$

$$NN_{akS} = \sum_{N} p^*_N X_{akN} + \sum_{N'} p^*_{N'} X_{akN'} + \sum_{N} q^*_N c_{ak} X_{akN} + \sum_{N'} q^*_{N'} c'_{ja} X_{akN'} + r^*\,[median_T\{X_{akT}\}]$$

We also remark that our methods relied on batch sequencing of general hematologic samples and may be subject to error in heavily biased cohorts that shift the median, in contrast to the use of a matched sample in the Sun study. Our methods were also independent of isoform, whereas the metric in the Sun study was optimized to exons 2-8 potentially biasing assessment of *KMT2A*-PTDs spanning different exons.

SNP analysis of NGS data and KASUMI6: SNP profiles were constructed over chromosome 11 for each of the cohorts and their respective assays (HSS, RHP, MYH) as follows. A master bedfile of effective coverage consisting of any genomic location of chromosome 11 with coverage $\geq$ 100 (HSS) or 50 (RHP, MYH) in at least one sample was first generated through "samtools depth". The intervals of this bedfile were then cross-referenced with gnomAD v2.1.1 exomes and genomes obtained through Google colab and its BigQuery interface into gnomAD. A master SNP bedfile was next constructed consisting of genomic positions having population allelic frequency $\geq$ 0.0001 for an ALT SNP (not indels), and these positions were subsequently evaluated for each sample through "samtools mpileup" to produce SNP profiles. A subset of these SNP genomic locations were subject to recurrent artifacts (e.g. from repetitive sequence, multi-mapping, etc), making estimation of allelic fractions unreliable. Suboptimal locations with a high empirical proportion of allelic fractions away from 0, 0.5, or 1 were identified and excluded. Other locations were subject to systematic bias for estimating the heterozygous state due to likely differential binding of probes or primers, and this was recognized through histograms and violin plots of empiric allelic fraction distributions. CN-LOH was assessed by manual review of the resulting SNP profiles, in conjunction with copy number profiles by BR-CNV, available karyotypic information, and empiric background distributions. SNP analysis of the KASUMI6 cell line was performed by downloading publicly available raw Cytoscan HD SNP array data (GSM4254134_Kasumi-6_CytoScanHD_Array_.CEL.gz) from Gene Expression Omnibus and re-processing it with the package Rawcopy (https://hub.docker.com/r/rawcopy/rawcopy; downloaded 08/30/2020) to obtain B-allele frequencies over chromosome 11 [4,5]. Cystoscan HD array SNP sites within candidate regions of CN-LOH in KASUMI6 were then queried in genotypes from the 1000 Genomes project (ALL.chr11.shapeit2_integrated_snvindels_v2a_27022019.GRCh38.phased.vcf.gz; downloaded 09/24/2020 from www.internationalgenome.org/data-portal) to determine a population distribution of heterozygous SNP counts over those sites.

Estimates of *KMT2A*-PTD allelic fractions from copy number levels: The allelic fraction (AF) of a *KMT2A*-PTD was defined as the ratio

AF = (copy number of *KMT2A*-PTD exons) / (copy number of distal *KMT2A* exons) - 1

In most circumstances, this ratio represented the bulk average of the number of *KMT2A*-PTD copies in a cell divided by the total number of wild-type and *KMT2A*-PTD copies, similar to the definition of an allelic fraction of a single nucleotide variant. For example, the AF of a cell with a *KMT2A*-PTD allele and a wild-type allele was 0.5 while a cell with a *KMT2A*-PTD allele that underwent CN-LOH was 1. In theory, this definition of AF could also exceed 1 due to copy number contributions that were not technically *KMT2A*-PTDs, for example episomal amplification or higher order linear replications (triplications, etc) producing the same breakpoint as a *KMT2A*-PTD, however the existence of these variants has not been reported before to our knowledge. Other structural variants involving *KMT2A* could also affect the AF, such as non-balanced rearrangements or those associated with gain, however these would generally not cause the AF to exceed 1 and would have different breakpoints from a *KMT2A*-PTD.

Split-read based detection, characterization, and allelic-fraction estimates of structural variants: Raw reads aligning to *KMT2A* were extracted from bamfiles generated by the standard pipelines prior to deduplication or UMI reduction, and realigned with bwa-mem v0.7.17 to hg19 for the case of HSS and MYH whereas RHP already used bwa-mem as its de facto aligner. Read pairs satisfying relatively stringent alignment criteria (concordant under bwa-mem with insertions totaling ≤2 bp, deletions totaling ≤2 bp, soft clips totaling ≤2 bp, and edit distance ≤5 bp in each read of a pair) were categorized as reference-reads and set aside for use in allelic-fraction estimates. Split-reads were defined as single ends of non-reference-reads having one (or rarely two) supplementary alignments under bwa-mem, and were each associated with a breakpoint signature consisting of a supplementary breakpoint, primary breakpoint, and overlap for every supplementary alignment, where the overlap could be positive (shared microhomology), negative (intervening unaligned sequence), or zero. Every breakpoint signature was further associated with a pair (or rarely triplet) of core CIGAR strings stripped of soft-clip and hard-clip components from the primary and secondary alignments. To minimize artifactual false positives, breakpoint signatures needed to either be (i) common to 3 or more split-reads with at least 3 different core CIGAR pairs or (ii) previously identified in the setting of a serial sample to undergo further consideration. Associated split-reads were next extended in-silico along reference chromosomes away from breakpoints to reach the nearest whole 1 kbp genomic coordinate at least 500 bp away from a breakpoint. Extended reads were then clustered into representatives referred to as structural variant mini-genomes, and the locations of mutant breakpoints along mini-genomes were determined from breakpoint signatures. Supporting mutant reads were identified by aligning non-reference reads directly to a mini-genome via bwa-mem and keeping paired-reads satisfying relatively stringent alignment criteria (concordant alignments extending 10 bp or more past a mutant breakpoint location in at least one read of the pair, and otherwise having insertions totaling ≤2 bp, deletions totaling ≤2 bp, soft clips totaling ≤2 bp, and edit distance ≤5 bp in each read of the pair). Mutant read counts, depth of coverage by genomic position, and breadth of coverage by genomic position, especially over the region bounded by mutant breakpoint(s) in the mini-genome, were used to evaluate the strength of evidence for a structural variant.

To estimate allelic fractions, supporting mutant reads were compared to reference reads that also extended 10 bp or more past a mutant breakpoint in at least one read of a pair, and additionally satisfied platform-specific criteria. For HC (MYH), applicable reference reads were further limited to sequenced fragments whose inferred centers were located proximal to the mutant breakpoint in order to exclude fragments that were more likely captured by baits past a mutant junction and thus not capable of hybridizing to a mutant genome. For AMP (HSS) and NEB (RHP), applicable reference reads were further limited to fragments derived from GSP2s (AMP) or baits (RHP) proximal to the mutant junction, which could be determined in these assays by alignment of the 5’ end of R2 (read 2). For general rearrangements, the allelic fraction was then calculated as [mutant reads / (mutant reads + reference reads)] similar to

SNVs. For *KMT2A*-PTDs, the allelic fraction was calculated as [mutant reads / reference reads] since PTD alleles were capable of generating mutant and reference reads. In particular, although PTD alleles initially appeared twice as likely in theory to generate fragments overlapping reference boundaries versus mutant junctions of a duplication, the platform specific criteria enabled proper accounting of supporting mutant reads and applicable reference reads.

RNA-sequencing analysis of *KMT2A*-PTDs in CCLE and clinical data: Compressed fasta files were downloaded from SRA by fastq-dump for select CCLE cell-lines: EOL1 (SRR8616218), KASUMI6 (SRR8615363), HL60 (SRR8616133), OCI-AML3 (SRR8615242), and KASUMI1 (SRR8615361). Paired-end reads containing certain exon:exon junctions of 30 bp length were identified and counted in the fasta files by "zgrep -B1" applied to the following sequences: (1) exon8:exon2 mutant junction (AAACCAAAAGAAAAGGATGAGCAATTCTTA and its reverse complement TAAGAATTGCTCATCCTTTTCTTTTGGTTT), (2) exon8:exon9 wild-type junction (AAACCAAAAGAAAAGGAAAAACCACCTCCG and its reverse complement CGGAGGTGGTTTTTCCTTTTCTTTTGGTTT), and (3) exon1:exon2 wild-type junction (GGCGGCAGCGGAGAGGATGAGCAATTCTTA and its reverse complement TAAGAATTGCTCATCCTCTCCGCTGCCGCC). Clinical RNA data (HFA) of *KMT2A*-PTDs in the MGH RNA/DNA cohort was assessed by identifying and counting split-read alignments in the HFA bamfiles (generated by bwa-mem) that corresponded to mutant or wild-type junctions and that originated from anchored primers capable of sequencing across the *KMT2A*-PTD mutant junction.

**Supplementary Figure Legends**

**Supp Fig 1.** Validation of BR-CNV estimates. Copy number estimates by BR-CNV of chromosome 21 in cases of AML of Down syndrome and various other genomic loci compared to clinical FISH data. Total copy number levels were obtained by integrating karyotype data to identify the diploid baseline.

**Supp Fig 2.** BR-CNV estimates of *KMT2A*-PTD exons. (A) Comparisons on 7 cases sequenced by both RHP and HSS with known *KMT2A*-PTDs identified by clinical RNA testing (HFA). Average levels of chromosome 15 targets were also compared, where 3 cases had trisomy 15 by karyotype. (B) Estimates by BR-CNV versus RobustCNV for RHP data, where RHP was the only assay with clinically validated copy number calling.

**Supp Fig 3.** *KMT2A*-PTDs with gain of 11q23.3 from the *KMT2A*-PTD allele. (A-B) P1 and P2 were described in Figure 1. (C) P3 progressed from MDS to AML with subsequent appearance of 11q23.3 gain from the *KMT2A*-PTD allele identified ~50 days before death. (D) P4 (AML) relapsed after transplant with appearance of 11q23.3 gain from the *KMT2A*-PTD allele and subsequent outgrowth of this clone prior to death. (E) Gain of 11q23.3 analysis for all serial samples from P1-P4. Analysis methods are provided in Figure 3. The final P3 time point was also described in Figure 3 as existing on an upper line corresponding to a near total mixture of complex *KMT2A*-PTD and standard simple *KMT2A*-PTD (with almost no wild-type). By contrast, P2 existed on a lower line corresponding to a complex *KMT2A*-PTD (with 2 gains of 11q23) at ~70% and wild type at ~30%, with no apparent simple *KMT2A*-PTD component.

**Supp Fig 4.** CN-LOH of *KMT2A*-PTDs from the oAML research cohort. (A-D) Four oAML cases (P5-P8) demonstrated *KMT2A*-PTDs with CN-LOH in the setting of normal (A, C), simple karyotypes (D), and unknown karyotypes (B). *KMT2A*-PTD in AML with normal cytogenetics has previously been thought to affect only a single allele (Caligiuri 1997). CN-LOH generally spanned the targeted and occasional off-target regions of 11q, where additional involvement of the 11p arm could not always be excluded The highest ratio (2.77) corresponded to 5.55 copies of *KMT2A* exons 2-8 with no other identified copy number changes; the gain could be entirely attributed to a single mutant junction consistent with a *KMT2A*-PTD since split-reads connecting intron 8 to intron 1 yielded a copy number estimate of 5.28 similar to the BR-CNV estimate of 5.55. This magnitude of gain in the context of a single mutant junction raised the possibility of a different mechanism such as episomal amplification. CN-LOH of 11q was also detected in 5 separate oAMLs harboring *CBL* mutations but not *KMT2A*-PTD, and 2 oAMLs with no identifiable 11q variants.

**Supp Fig 5.** CN-LOH of *KMT2A*-PTDs from the clinical cohorts. (A-G) Seven cases (P9-P15) tested by the clinical NGS assays demonstrated *KMT2A*-PTDs with CN-LOH and were associated with normal (2), simple (4), and unknown karyotypes (1). The cases were comprised of 4 MDS (P9-P11, P15) and one PV (P12) with secondary AML transformation, one MDS (P13) without progression, and one myeloid sarcoma (P14). Due to limited targeted coverage of 11q in the clinical assays, a few of these cases had informative SNPs adjacent to *KMT2A* but not spanning both sides; nevertheless, involvement of the *KMT2A* locus was inferred as the explanation of high copy number gain. SNP analysis was occasionally complicated by transplant status but could be resolved in the context of low donor chimerism (P11).

**Supp Fig 6.** Evolution of *KMT2A*-PTD and co-mutations across serial samples. (A) In P3, *KMT2A*-PTD complexity (as described in Supp Fig 3) was the only identifiable clonal event tracking the evolution of blasts at the final time point of progression. Distal gain complexity was not detectable by bulk NGS at the intermediate time point of AML diagnosis, however its subclonal presence at AML diagnosis could be deduced from clonal hierarchy. Namely, an *NRAS* hotspot variant was inferred as subclonal to the complex *KMT2A*-PTD based on initial absence at the MDS time point and the estimated clone sizes at the final time point (70% cells with complex *KMT2A*-PTD, 30% cells with simple *KMT2A*-PTD, and 35% cells with *NRAS* corresponding to 17.5% VAF) (see also Figure 3). Thus, the emergence of the *NRAS* variant at AML diagnosis (1.4% VAF) implied the presence of *KMT2A*-PTD complexity below the limit of NGS detection. (B) In P4, an initially subclonal *NF1* loss-of-functon SNV grew out as the dominant clone at relapse. The initial timepoint also had single copy loss of *NF1* at a relatively high inferred allelic fraction, thus the *NF1* SNV was likely already associated with LOH at the initial timepoint. *KMT2A*-PTD complexity also emerged at relapse. Estimated percent tumor cells involved by *KMT2A*-PTD complexity after relapse (48%, 71%, and 87%) suggested a subclonal relationship to the biallelic *NF1* alterations (61%, 76%, and 88%) with relative outgrowth of the subclone. (C) In P11, a *RUNX1* variant with CN-LOH was newly detected and the dominant clone upon MDS relapse and expanded during AML progression. A simple *KMT2A*-PTD was present at the initial MDS time point prior to relapse, and experienced CN-LOH during tumor evolution, however a robust CN-LOH signal was only possible at the final time point (AML) due to earlier chimeric states from transplant. Estimated percentage of involved tumor cells was again less for the complex *KMT2A*-PTD event (88%) compared to *RUNX1* (96%) at the final time point, suggesting an evolutionary timeline in the predominant clone of a parental simple *KMT2A*-PTD followed by *RUNX1* SNV and CN-LOH events, followed by *KMT2A*-PTD CN-LOH.

**Supp Fig 7.** Copy number ratios of *KMT2A*-PTDs by isoform at first available time point (A) and at maximum burden time point (B). High ratios indicative of complexity were most prevalent for e8e2, while also present in a smaller cluster for e10e2.

**Supp Fig 8.** Correlation of *KMT2A*-PTD RNA expression to DNA allelic burden. The MGH RNA/DNA cohort contained 30 *KMT2A*-PTD samples from 20 patients with concurrent HSS and HFA clinical testing, while the cell lines KASUMI6 and EOL1 had CCLE public data from whole-transcriptome RNA-seq and Affymetrix Genome-Wide Human SNP Array 6.0. RNA reads containing the mutant splice junction were normalized relative to reads containing reference splice junctions, which could be derived from mutant or wild-type transcripts since tandem duplications contained both mutant and wild-type junctions. AMP: anchored multiplex PCR used by the HFA and HSS assays. WT: wild-type. SR: split-read. VAF: variant allele fraction.

**Supp Fig 9.** Incidence of *KMT2A*-PTD in an unselected cohort of new diagnoses (BWH 2019-2020). AA: aplastic anemia, AML: acute myeloid leukemia, B-ALL: B-cell acute lymphoblastic leukemia, BPCDN: blastic plasmacytoid dendritic cell neoplasm, CNL: chronic neutrophlic leukemia, CML: chronic myeloid leukemia, ET: essential thrombocytopenia, HCL: hairy cell leukemia, HLH: hemophagocytic lymphohistiocytosis, LGL: large granula lymphocytic leukemia, LYMPH: lymphoma, MAST: mastocytosis, MDS: myelodysplastic syndrome, MM: multiple myeloma, MPN: myeloproliferative neoplasm (other), PMF: primary myelofibrosis, PV: polycythemia vera, T-ALL: T-cell acute lymphoblastic leukemia.

**Supp Fig 10.** Split-read based estimates of structural variant and *KMT2A*-PTD allelic fractions. (A) Split-read based estimates of *KMT2A* rearrangements compared to concurrent clinical *KMT2A* FISH. As a side benefit from this effort, *KMT2A* split-reads revealed breakpoints of several novel (non-PTD) *KMT2A* rearrangements of unknown significance (*KMT2A-AHCYL2*, *LINC01531-KMT2A,* *MECR-KMT2A*, and *KMT2A-GATAD2*), and demonstrated a sensitivity of 55% (6/11) and specificity of 100% (5/5) relative to cytogenetics in one of the assays (RHP) despite not explicitly targeting *KMT2A* introns. (B) Split-read versus BR-CNV estimates of *KMT2A*-PTD copy number ratios, restricted to cases detected by both BR-CNV and split-read analysis with coverage depth above 200 around the breakpoints (split-reads + reference reads). As described earlier, BR-CNV estimates correlated with FISH over various loci and was thus extrapolated to perform well across the targeted genome including *KMT2A*.

**Supp Fig 11.** *DDX41*-related AML with *KMT2A*-PTDs assessed by split-reads. (A) A novel *KMT2A*-PTD involving exons 2-15 was present as a minor subclone in a case of *DDX41*-related AML, with split-read based allelic fraction estimates of 1.0% (15 split-reads / 1484 reference), 3.0% (69/2327), and 2.1% (53/2481), contrary to typical clonal *KMT2A*-PTDs that are considered critical to AML development. Clinical RNA testing also revealed aberrant expression of the e15e2 isoform. (B) Split-reads from an intronic area with low coverage yielded inaccurate estimates for a *KMT2A*-PTD involving exons 2-10 from a second *DDX41*-related AML but were still present at an MRD timepoint.

**Supp Fig 12.** Split-reads help resolve potentially ambiguous copy number signals associated with pitfalls and unusual *KMT2A*-PTD cases. A common challenge occurred when partial *KMT2A* copy number gain included exon 1, which could represent multiple possibilities: (A-B) a non-standard *KMT2A*-PTD spanning exons 1-8 with split-reads confirming genomic breakpoints in intron 8 and immediately proximal to exon 1, and (C-D) *KMT2A* rearrangement, especially *KMT2A-MLLT10*, where partial duplication of the 5’ segment of *KMT2A* is known to occur rarely but recurrently (Jarosova 2005: 16213369, Sarova 2010: 20471515, Fukushima 2017: 29445560) and was seen in a case from our oAML cohort with gain of *KMT2A* exons 1-6 by BR-CNV and evidence of *KMT2A-MLLT10* by both karyotype 49XY,+6,+8,+19,der(10)ins(10;11)(p13;q23.3q23.3) and explicit split-read capture of genomic *KMT2A-MLLT10* breakpoints (chr11:118351187, chr10:21985531; insG). Not shown: mischaracterization of the copy number level of exon 1 relative to the true genomic breakpoints is another common challenge due to high GC content of exon 1 and may have occurred in cases of the MGH RNA/DNA cohort with discordant *KMT2A*-PTD boundaries between BR-CNV and RNA (e1e8 instead of e2e8 in 1 case, e1e10 instead of e2e10 in 2 cases, and e3e9 instead of e2e10 in the lone FFPE case). (E-F) It was also important to incorporate the overall level of chromosome 11 in copy number based assessment for *KMT2A*-PTDs, since relative gain of 5’ *KMT2A* in the context of bulk loss of 3’ *KMT2A* was more likely to represent a *KMT2A* rearrangement, including multiple examples from our cohorts. Thus, algorithms in the literature that only consider exonic coverage ratios may mis-identify these examples as *KMT2A*-PTDs. Not shown: Although extremely rare, copy number gains not including exon 1 may potentially represent false positives; namely, predominant *KMT2A-AF9* expression has been reported from non-tandem duplication of *KMT2A* exons 2-8 interrupted by insertion of 3’ *AF9*, albeit in a B-ALL case and not a myeloid malignancy (Whitman 2001: 11196198). (G-H) Finally, integration of split-reads and copy number data showed promise at characterizing rare difficult cases, including an oAML case where split-reads involving *KMT2A* intron 8 and chromosome 4 were insufficient to explain the magnitude of copy number gain of *KMT2A* exons 2-8. *KMT2A* exon 1 demonstrated gain to a lesser magnitude than exons 2-8, raising the possibility of a separate *KMT2A*-PTD.

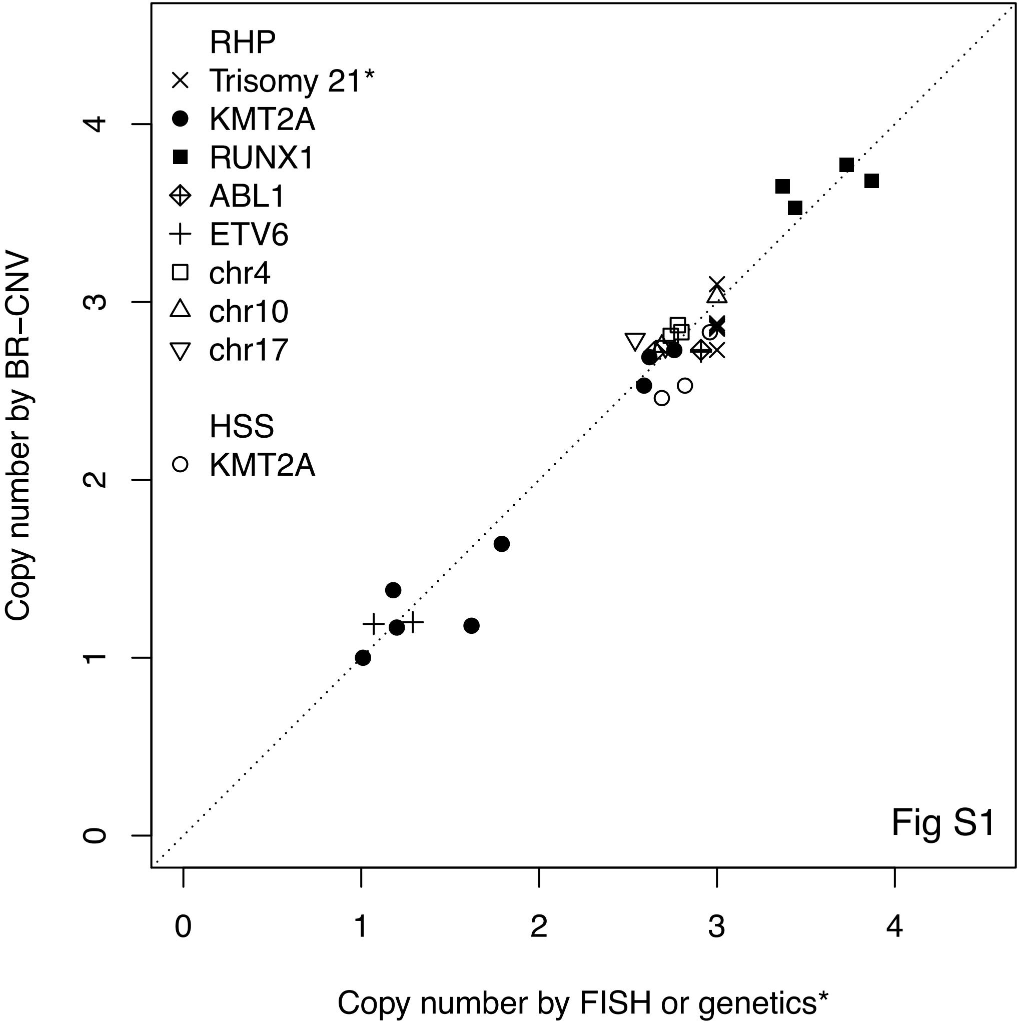
RHP
Trisomy 21*
KMT2A
RUNX1
ABL1
ETV6
chr4
chr10
chr17
HSS
KMT2A
Copy number by BR−CNV
Copy number by FISH or genetics*
0
1
2
3
4
Fig S1

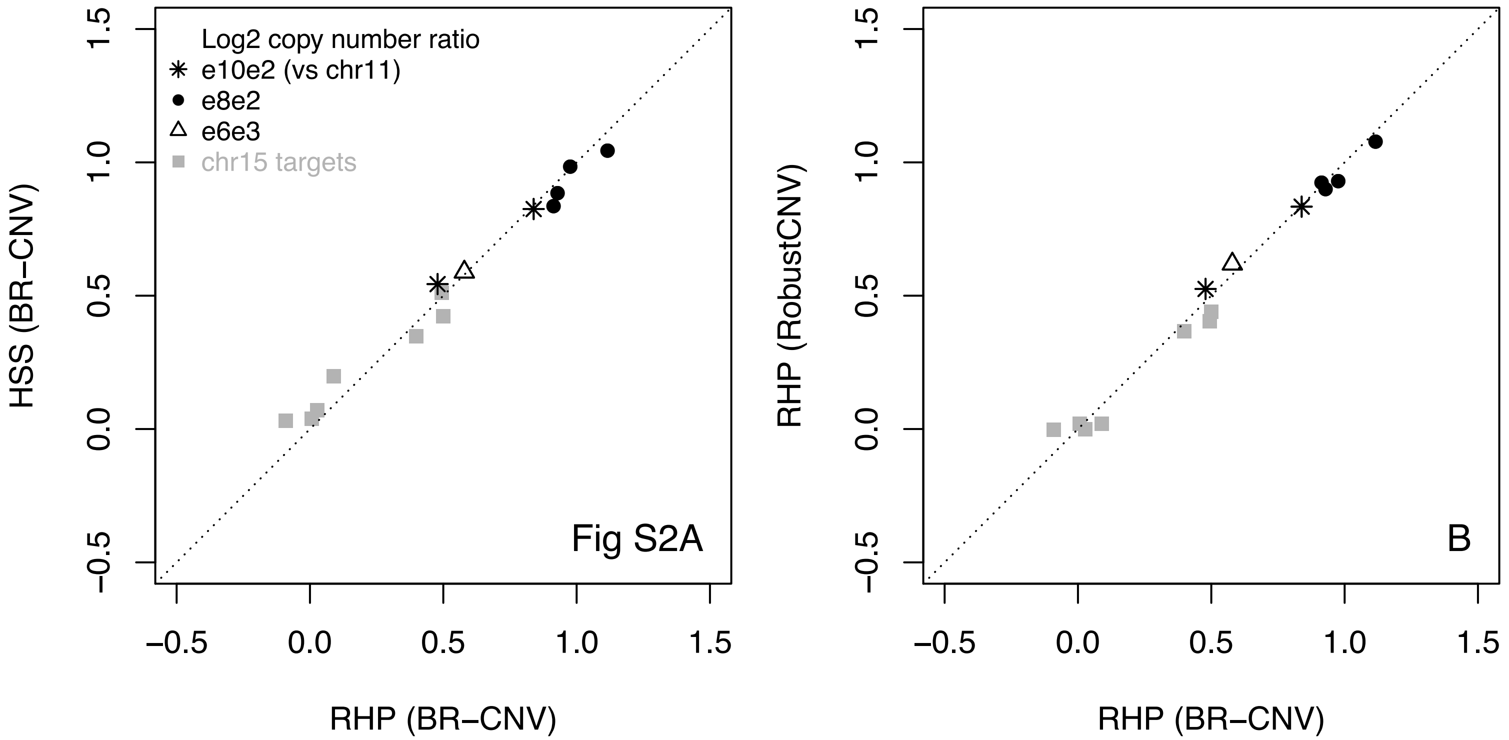

Log2 copy number ratio
e10e2 (vs chr11)
e8e2
e6e3
chr15 targets
Fig S2A
B
HSS (BR−CNV)
RHP (RobustCNV)
RHP (BR−CNV)
RHP (BR−CNV)

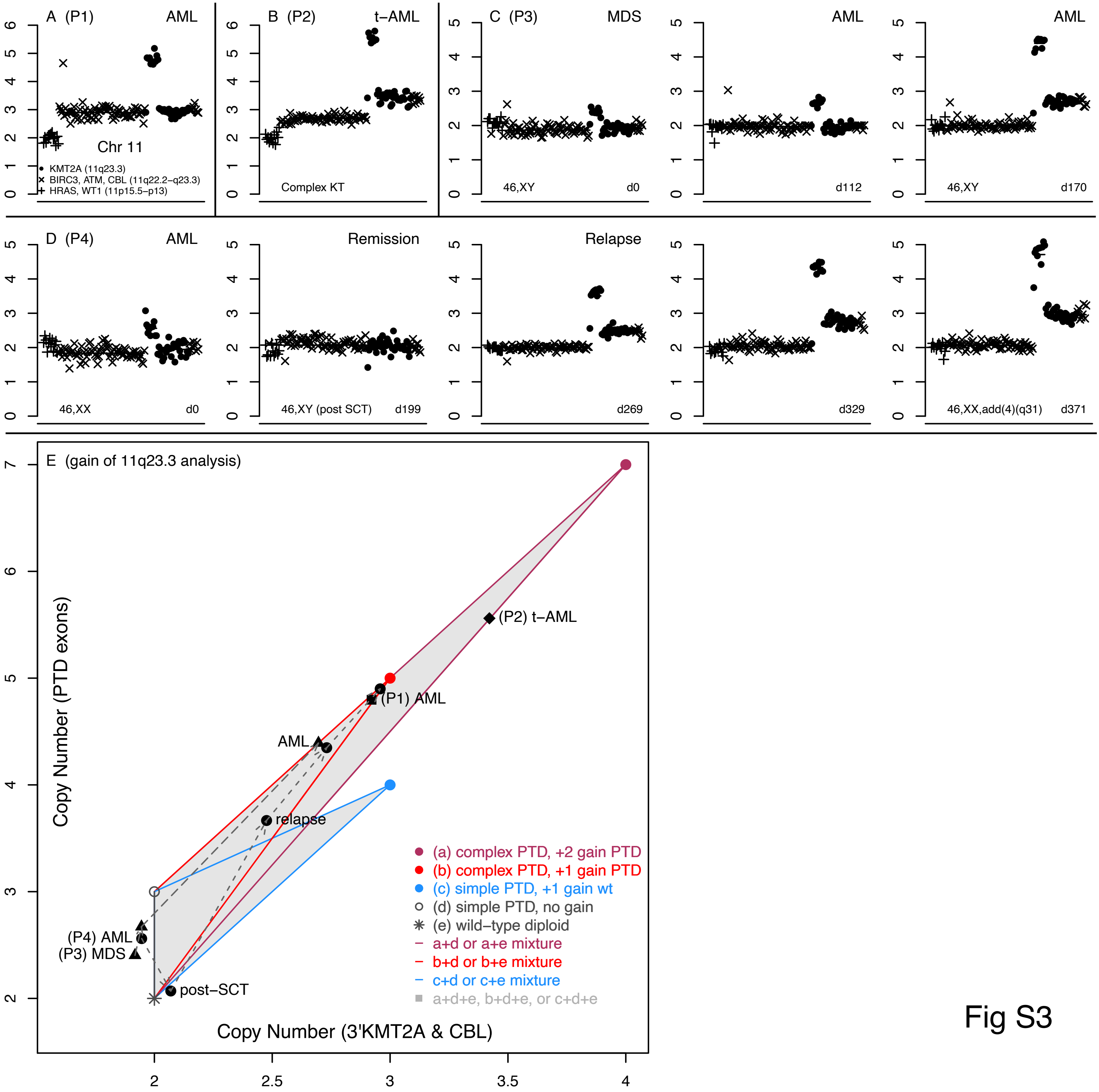

A (P1)
AML
Chr 11
KMT2A (11q23.3)
BIRC3, ATM, CBL (11q22.2–q23.3)
HRAS, WT1 (11p15.5–p13)
B (P2)
t–AML
Complex KT
C (P3)
MDS
46,XY
d0
AML
d112
AML
46,XY
d170
D (P4)
AML
46,XX
d0
Remission
46,XY (post SCT)
d199
Relapse
d269
d329
46,XX,add(4)(q31)
d371
E (gain of 11q23.3 analysis)
Copy Number (PTD exons)
Copy Number (3'KMT2A & CBL)
(P2) t–AML
(P1) AML
AML
relapse
(P4) AML
(P3) MDS
post–SCT
(a) complex PTD, +2 gain PTD
(b) complex PTD, +1 gain PTD
(c) simple PTD, +1 gain wt
(d) simple PTD, no gain
(e) wild–type diploid
a+d or a+e mixture
b+d or b+e mixture
c+d or c+e mixture
a+d+e, b+d+e, or c+d+e


Fig S3

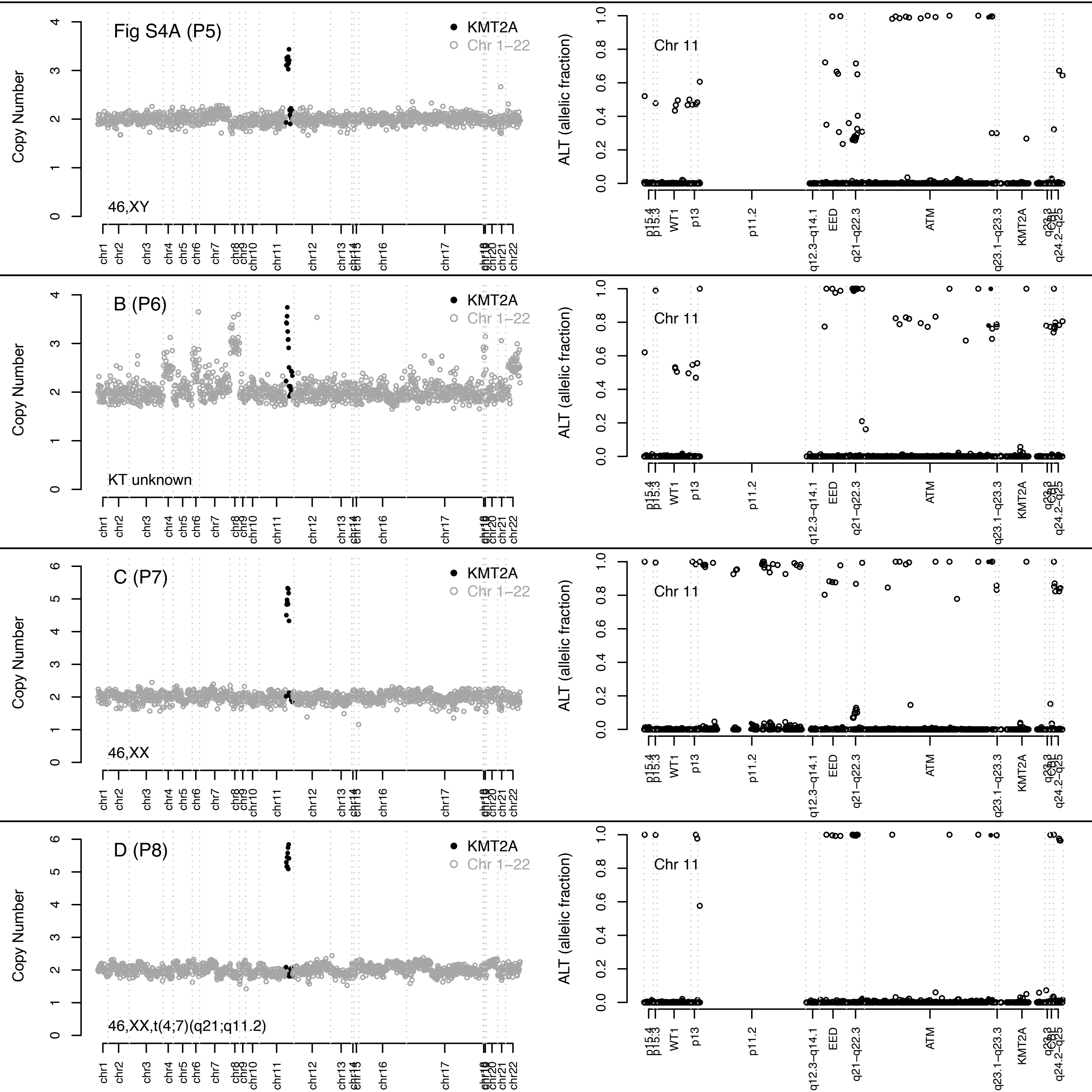

Fig S4A (P5)
46,XY
B (P6)
KT unknown
C (P7)
46,XX
D (P8)
46,XX,t(4;7)(q21;q11.2)
KMT2A
Chr 1–22
Copy Number
ALT (allelic fraction)
Chr 11
chr1
chr2
chr3
chr4
chr5
chr6
chr7
chr8
chr9
chr10
chr11
chr12
chr13
chr14
chr15
chr16
chr17
chr18
chr19
chr20
chr21
chr22
p15.4
p15.3
WT1
p13
p11.2
q12.3–q14.1
EED
q21–q22.3
ATM
q23.1–q23.3
KMT2A
q23.3
q24.2–q25

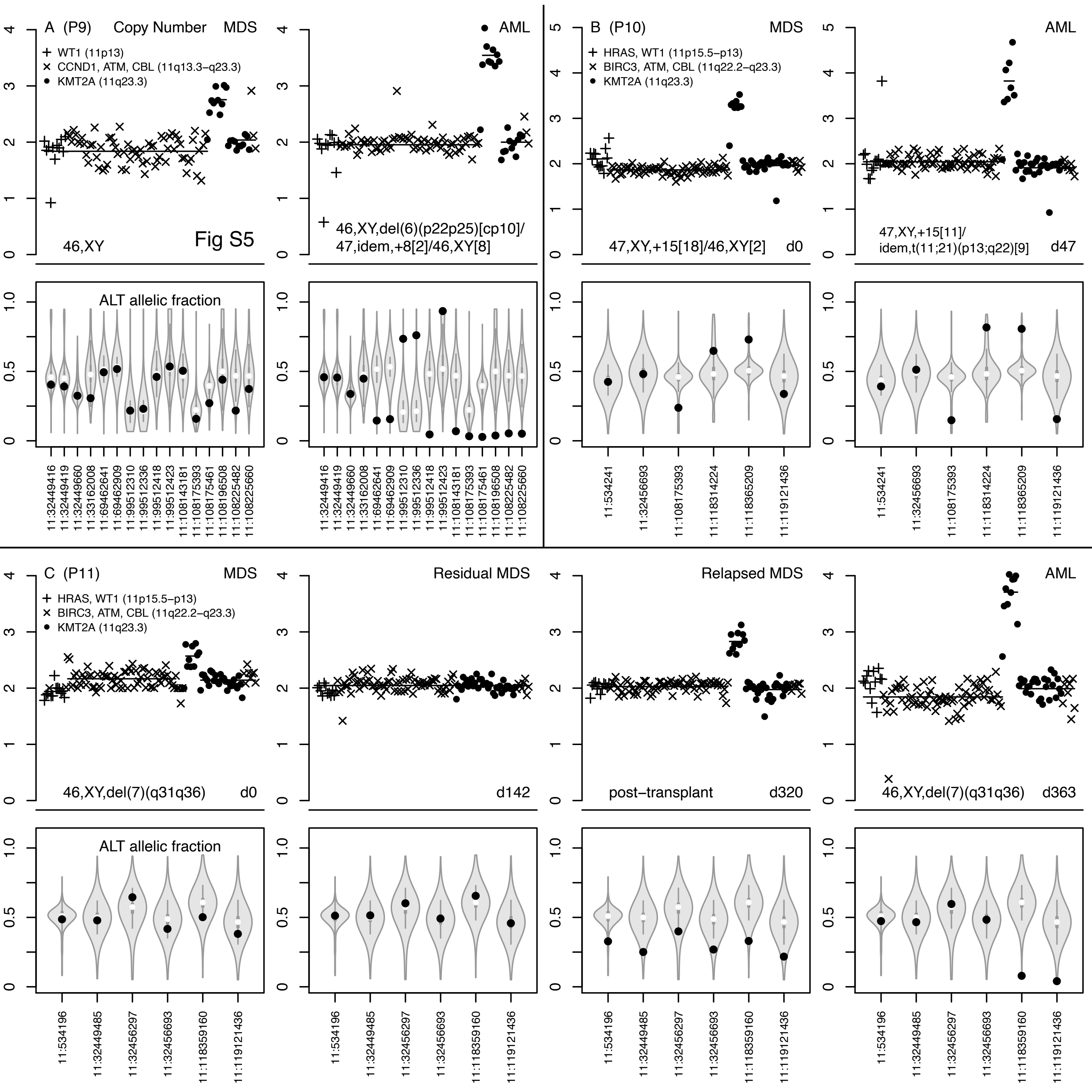

A (P9) Copy Number MDS
WT1 (11p13)
CCND1, ATM, CBL (11q13.3−q23.3)
KMT2A (11q23.3)
46,XY
Fig S5
AML
46,XY,del(6)(p22p25)[cp10]/
47,idem,+8[2]/46,XY[8]
B (P10) MDS
HRAS, WT1 (11p15.5−p13)
BIRC3, ATM, CBL (11q22.2−q23.3)
KMT2A (11q23.3)
47,XY,+15[18]/46,XY[2]
d0
AML
47,XY,+15[11]/
idem,t(11;21)(p13;q22)[9]
d47
ALT allelic fraction
11:32449416
11:32449419
11:32449660
11:33162008
11:69462641
11:69462909
11:99512310
11:99512336
11:99512418
11:99512423
11:108143181
11:108175393
11:108175461
11:108196508
11:108225482
11:108225660
11:534241
11:32456693
11:108175393
11:118314224
11:118365209
11:119121436
C (P11) MDS
HRAS, WT1 (11p15.5−p13)
BIRC3, ATM, CBL (11q22.2−q23.3)
KMT2A (11q23.3)
46,XY,del(7)(q31q36)
d0
Residual MDS
d142
Relapsed MDS
post−transplant
d320
AML
46,XY,del(7)(q31q36)
d363
ALT allelic fraction
11:534196
11:32449485
11:32456297
11:32456693
11:118359160
11:119121436

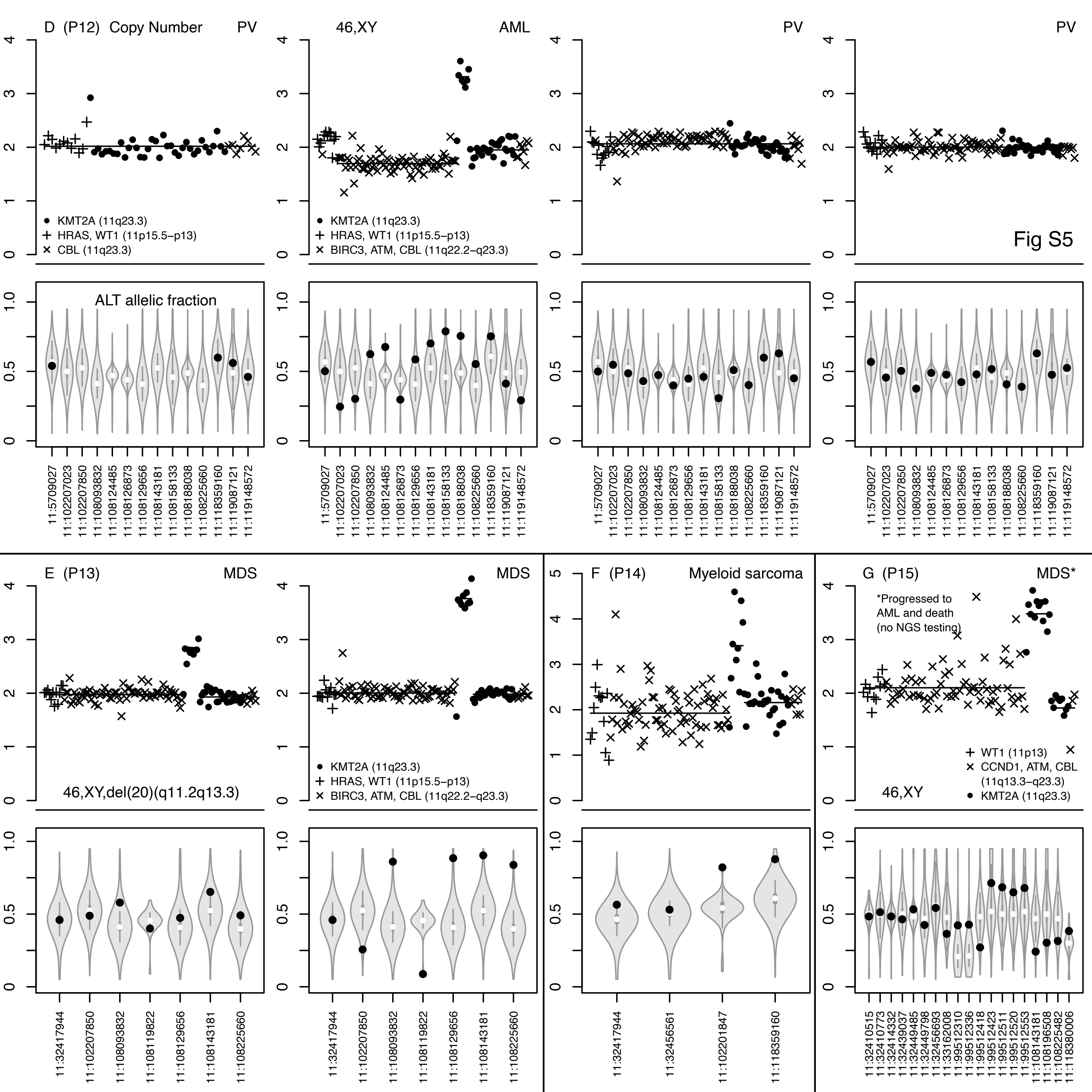
Fig S5
D (P12) Copy Number
PV
46,XY
AML
PV
PV
KMT2A (11q23.3)
HRAS, WT1 (11p15.5−p13)
CBL (11q23.3)
KMT2A (11q23.3)
HRAS, WT1 (11p15.5−p13)
BIRC3, ATM, CBL (11q22.2−q23.3)
ALT allelic fraction
11:5709027
11:102207023
11:102207850
11:108093832
11:108124485
11:108126873
11:108129656
11:108143181
11:108158133
11:108188038
11:108225660
11:118359160
11:119087121
11:119148572
E (P13)
MDS
MDS
46,XY,del(20)(q11.2q13.3)
KMT2A (11q23.3)
HRAS, WT1 (11p15.5−p13)
BIRC3, ATM, CBL (11q22.2−q23.3)
11:32417944
11:102207850
11:108093832
11:108119822
11:108129656
11:108143181
11:108225660
F (P14)
Myeloid sarcoma
11:32417944
11:32456561
11:102201847
11:118359160
G (P15)
MDS*
*Progressed to AML and death (no NGS testing)
WT1 (11p13)
CCND1, ATM, CBL (11q13.3−q23.3)
KMT2A (11q23.3)
46,XY
11:32410515
11:32410773
11:32414332
11:32439037
11:32449485
11:32449798
11:32456693
11:33162008
11:99512310
11:99512336
11:99512418
11:99512423
11:99512511
11:99512520
11:99512553
11:108143181
11:108196508
11:108225482
11:118380006

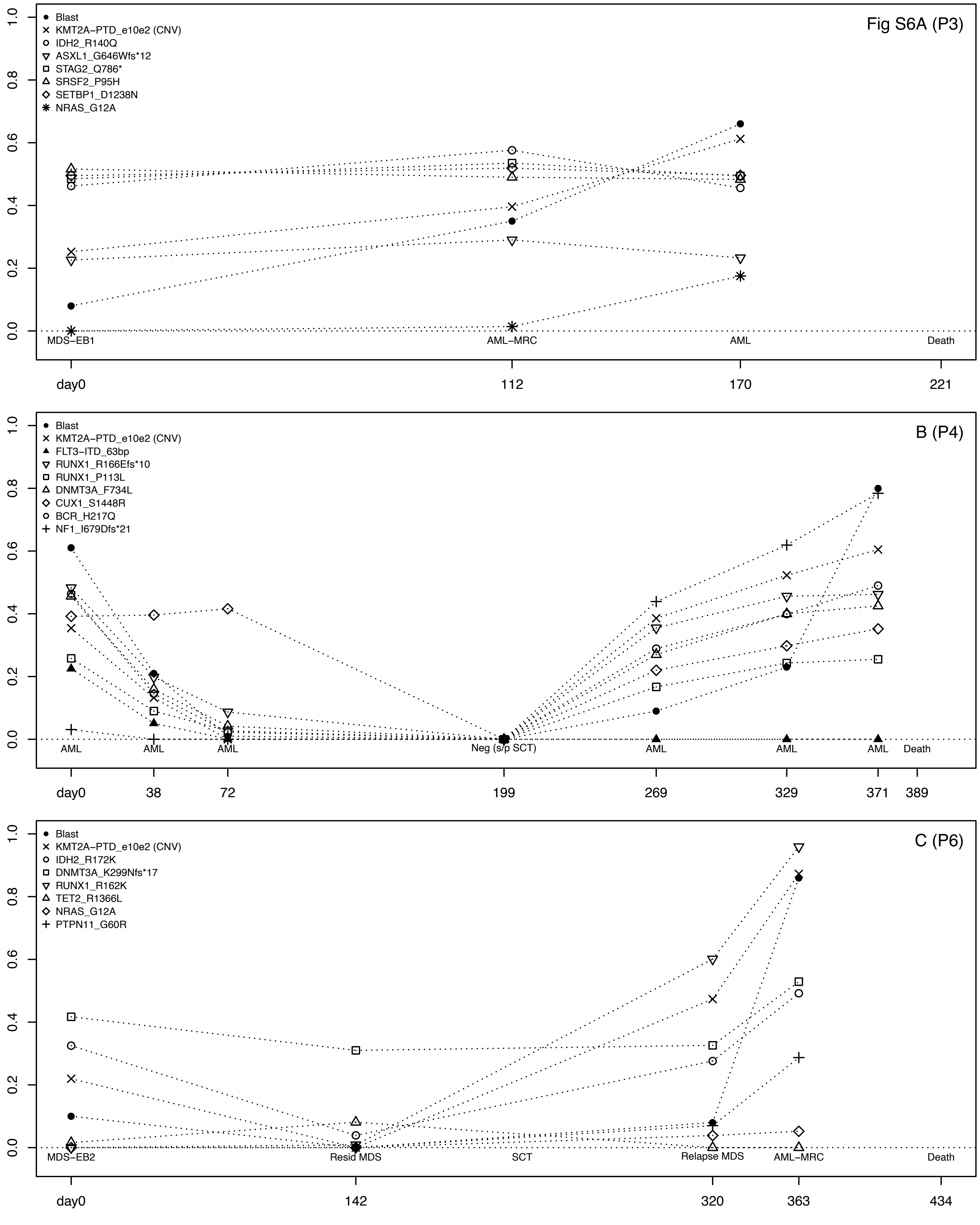
Fig S6A (P3)
Blast
KMT2A−PTD_e10e2 (CNV)
IDH2_R140Q
ASXL1_G646Wfs*12
STAG2_Q786*
SRSF2_P95H
SETBP1_D1238N
NRAS_G12A
MDS−EB1
AML−MRC
AML
Death
day0
112
170
221
B (P4)
Blast
KMT2A−PTD_e10e2 (CNV)
FLT3−ITD_63bp
RUNX1_R166Efs*10
RUNX1_P113L
DNMT3A_F734L
CUX1_S1448R
BCR_H217Q
NF1_I679Dfs*21
AML
Neg (s/p SCT)
Death
day0
38
72
199
269
329
371
389
C (P6)
Blast
KMT2A−PTD_e10e2 (CNV)
IDH2_R172K
DNMT3A_K299Nfs*17
RUNX1_R162K
TET2_R1366L
NRAS_G12A
PTPN11_G60R
MDS−EB2
Resid MDS
SCT
Relapse MDS
AML−MRC
Death
day0
142
320
363
434

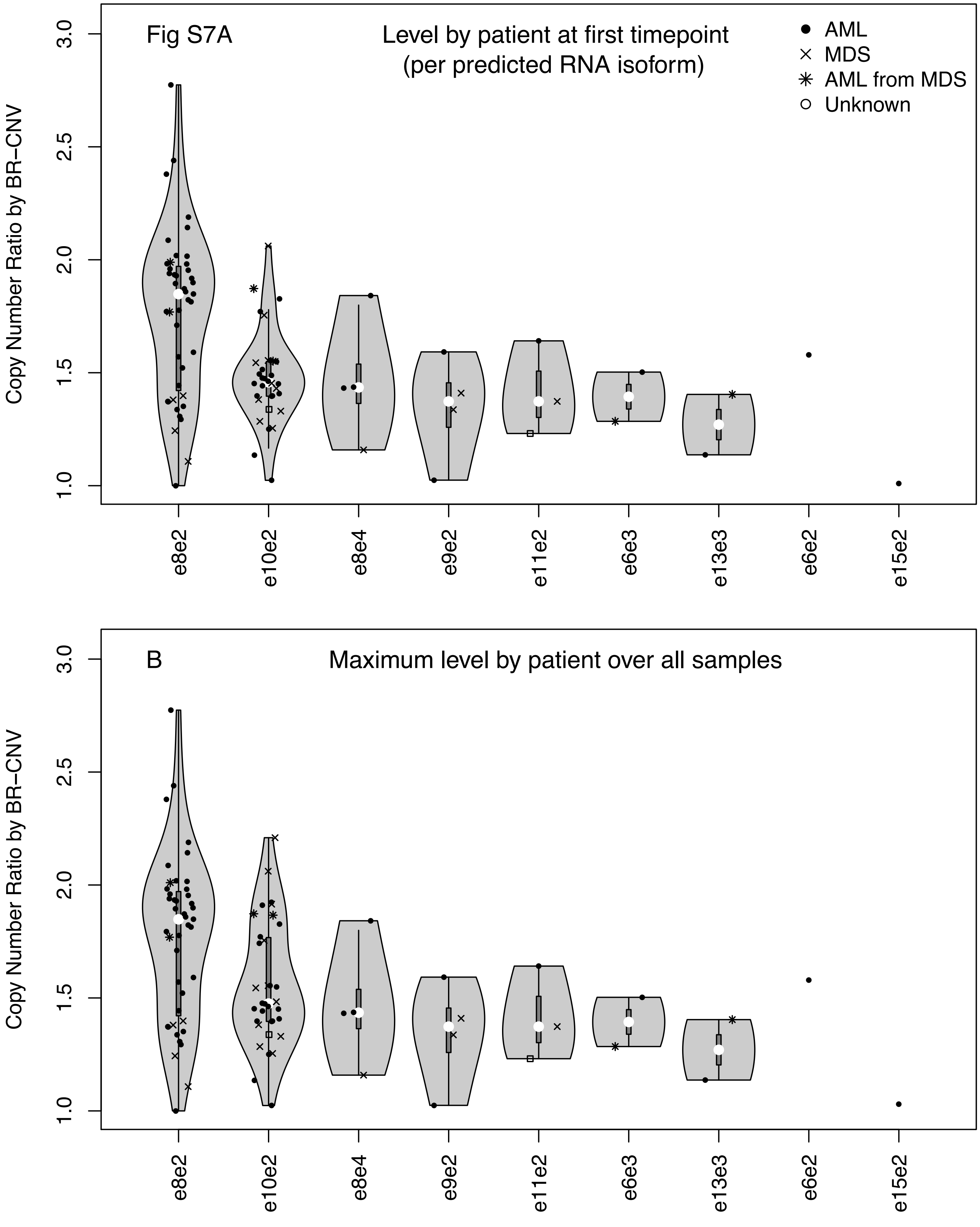
Fig S7A
Level by patient at first timepoint
(per predicted RNA isoform)
AML
MDS
AML from MDS
Unknown
Copy Number Ratio by BR−CNV
1.0
1.5
2.0
2.5
3.0
e8e2
e10e2
e8e4
e9e2
e11e2
e6e3
e13e3
e6e2
e15e2
B
Maximum level by patient over all samples
Copy Number Ratio by BR−CNV
1.0
1.5
2.0
2.5
3.0
e8e2
e10e2
e8e4
e9e2
e11e2
e6e3
e13e3
e6e2
e15e2

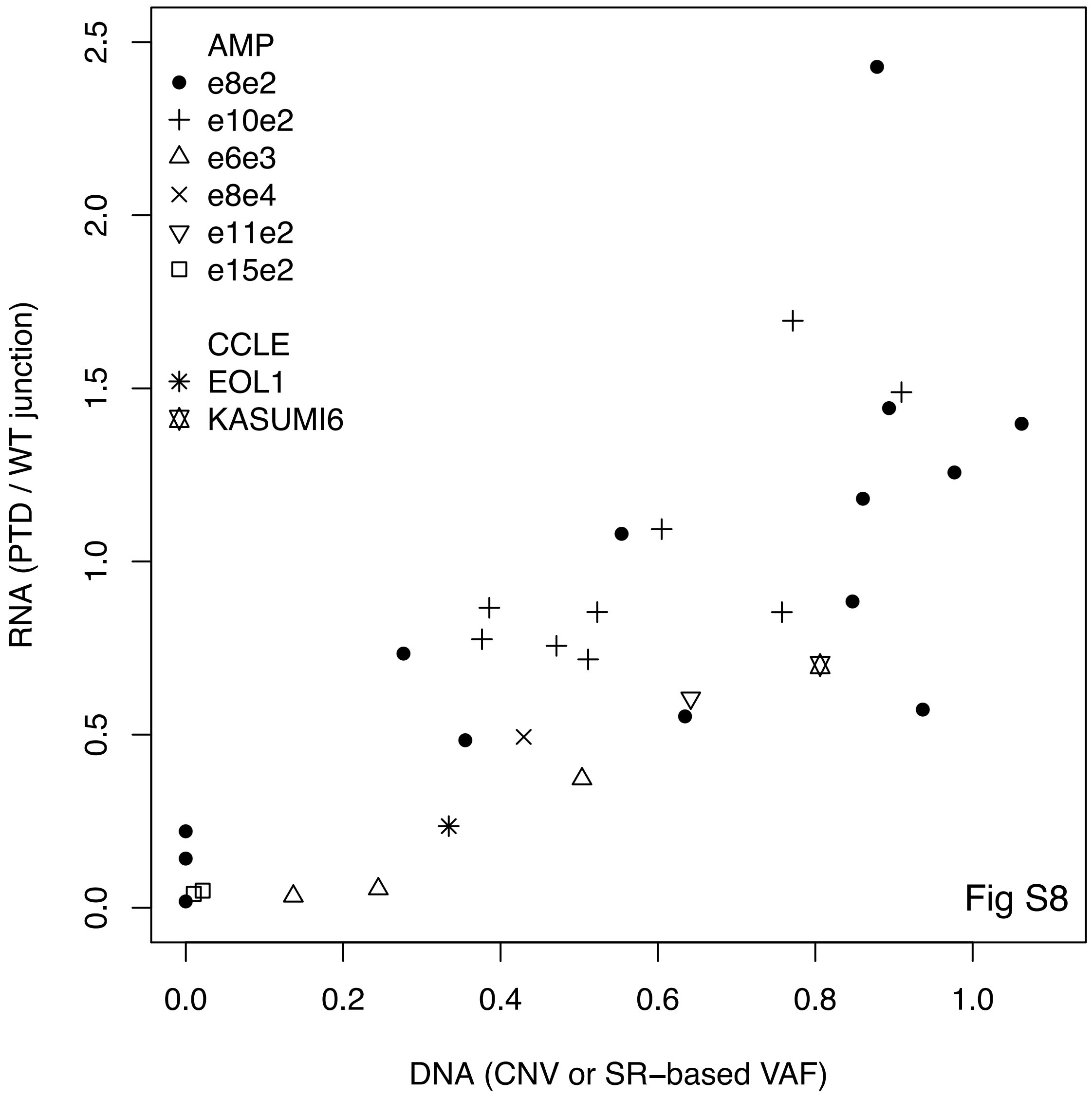
AMP
e8e2
e10e2
e6e3
e8e4
e11e2
e15e2
CCLE
EOL1
KASUMI6
Fig S8
RNA (PTD / WT junction)
DNA (CNV or SR−based VAF)
0.0
0.5
1.0
1.5
2.0
2.5
0.0
0.2
0.4
0.6
0.8
1.0

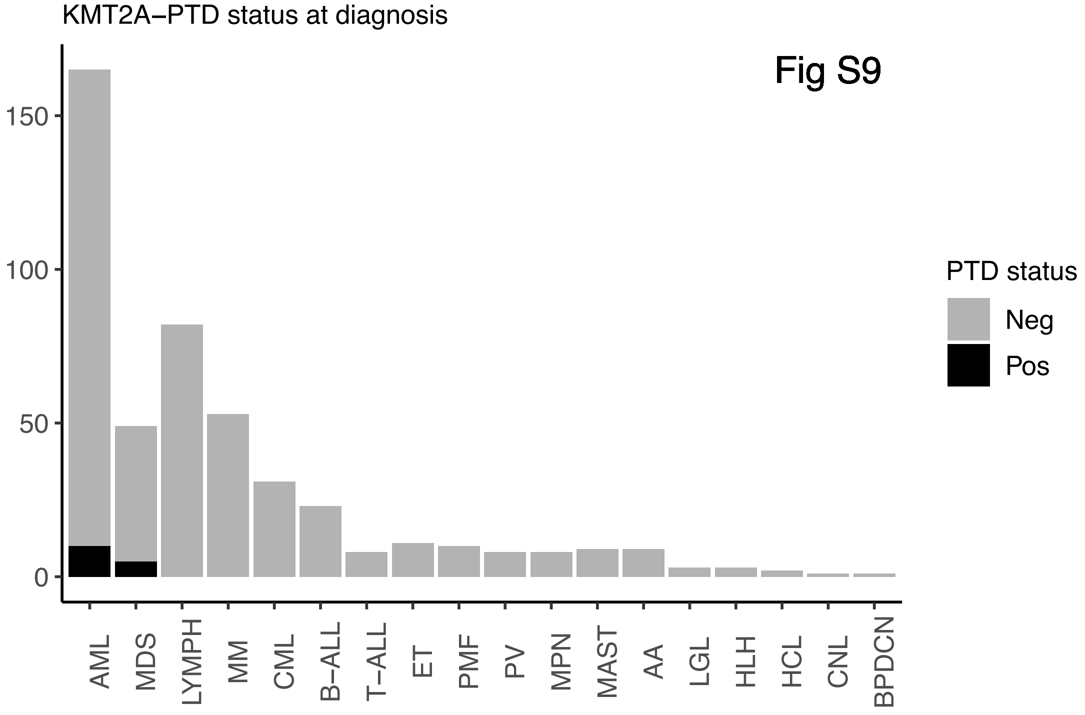

KMT2A−PTD status at diagnosis
Fig S9
150
100
50
0
AML
MDS
LYMPH
MM
CML
B−ALL
T−ALL
ET
PMF
PV
MPN
MAST
AA
LGL
HLH
HCL
CNL
BPDCN
PTD status
Neg
Pos

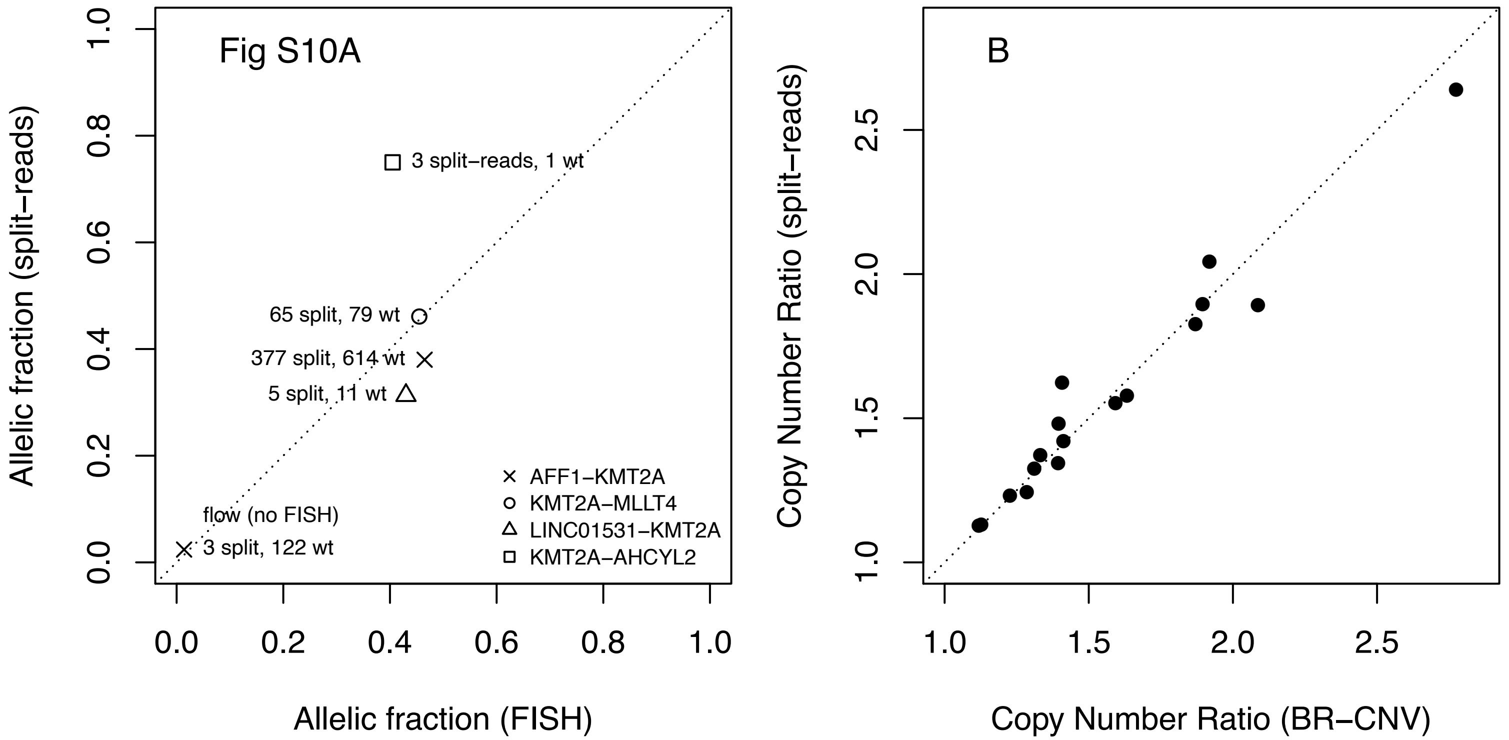
Fig S10A
3 split−reads, 1 wt
65 split, 79 wt
377 split, 614 wt
5 split, 11 wt
flow (no FISH)
3 split, 122 wt
AFF1−KMT2A
KMT2A−MLLT4
LINC01531−KMT2A
KMT2A−AHCYL2
Allelic fraction (split−reads)
Allelic fraction (FISH)
0.0
0.2
0.4
0.6
0.8
1.0
B
Copy Number Ratio (split−reads)
Copy Number Ratio (BR−CNV)
1.0
1.5
2.0
2.5

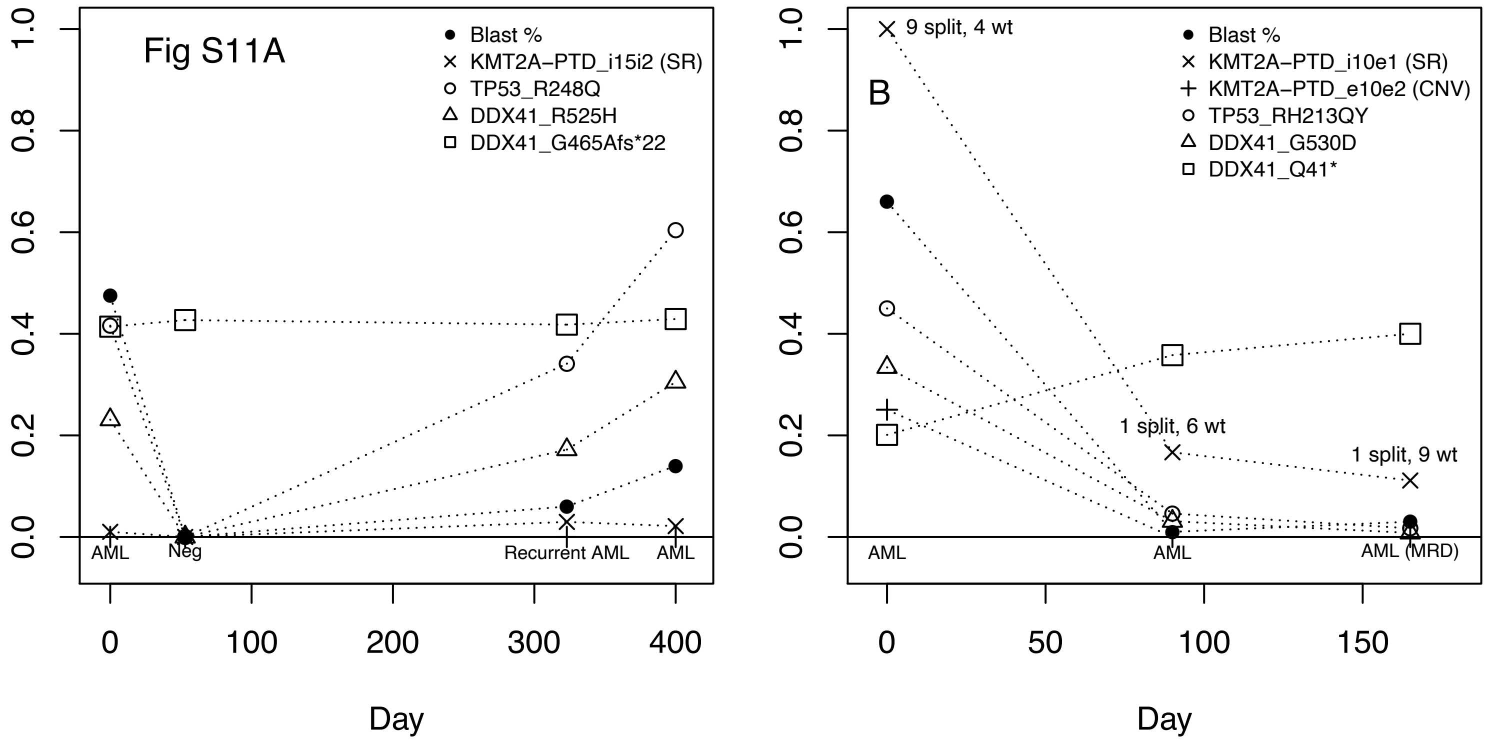
Fig S11A
Blast %
KMT2A−PTD_i15i2 (SR)
TP53_R248Q
DDX41_R525H
DDX41_G465Afs*22
AML
Neg
Recurrent AML
AML
Day
B
9 split, 4 wt
Blast %
KMT2A−PTD_i10e1 (SR)
KMT2A−PTD_e10e2 (CNV)
TP53_RH213QY
DDX41_G530D
DDX41_Q41*
1 split, 6 wt
1 split, 9 wt
AML
AML
AML (MRD)
Day

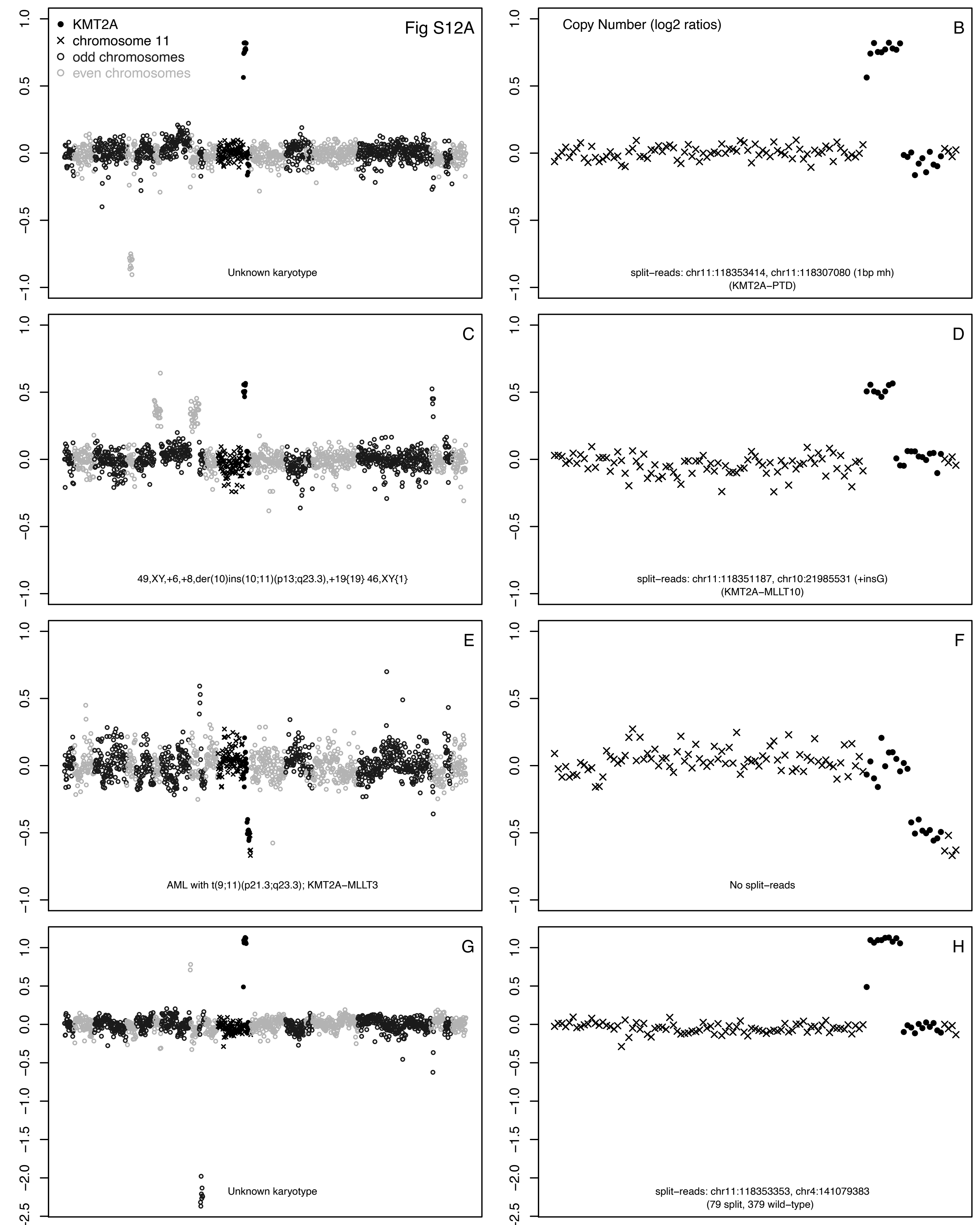

Fig S12A
KMT2A
chromosome 11
odd chromosomes
even chromosomes
Unknown karyotype
Copy Number (log2 ratios)
B
split−reads: chr11:118353414, chr11:118307080 (1bp mh)
(KMT2A−PTD)
C
49,XY,+6,+8,der(10)ins(10;11)(p13;q23.3),+19{19} 46,XY{1}
D
split−reads: chr11:118351187, chr10:21985531 (+insG)
(KMT2A−MLLT10)
E
AML with t(9;11)(p21.3;q23.3); KMT2A−MLLT3
F
No split−reads
G
Unknown karyotype
H
split−reads: chr11:118353353, chr4:141079383
(79 split, 379 wild−type)
